# Real-time Dissection and Forecast of Infection Dynamics during a Pandemic

**DOI:** 10.1101/2023.03.02.23286502

**Authors:** Steven Schulz, Richard Pastor, Cenk Koyuncuoglu, Forrest W. Crawford, Detlef Zernick, André Karch, Sten Rüdiger

## Abstract

Pandemic preparedness requires institutions, including public health authorities and governments, to detect, survey and control outbreaks. To maintain an accurate, quantitative and up-to-date picture of an epidemic crisis is key. For SARS-CoV-2, this was mostly achieved by ascertaining incidence numbers and the effective reproductive number (*R*_eff_), which counts how many people an infected person is likely to infect on average. These numbers give strong hints on past infection dynamics in a population but fail to clearly characterize current and future dynamics as well as potential effects of pharmaceutical and non-pharmaceutical interventions. We show that, by using and combining infection surveillance and population-scale contact statistics, we can obtain a better understanding of the drivers of epidemic waves and the effectiveness of interventions. This approach can provide a real-time picture, thus saving not only many lives by quickly allowing adaptation of the health policies but also alleviating economic and other burdens if an intervention proves ineffective. We factorize *R*_eff_ into contacts and relative transmissibility: Both signals can be used, individually and combined, to identify driving forces of an epidemic, monitoring and assessing interventions, as well as projecting an epidemic’s future trajectory. Using data for SARS-CoV-2 and Influenza from 2019 onward in Germany, we provide evidence for the usefulness of our approach. In particular, we find that the effects from physical distancing and lockdowns as well as vaccination campaigns are dominant.

## 1. Introduction

Infectious diseases represent serious threats to an ever increasingly connected humankind, on par with e.g. natural disasters and infrastructure failures. Epidemic preparedness – the ability to predict and mitigate future epidemic outbreaks – has thus risen to one of the most pressing challenges in modern societies and recently focused a wealth of research efforts building on a variety of data [1] in response to awareness elicited by the SARS-CoV-2 pandemic [2].

pidemic dynamics are shaped at the crossroads of human and viral driving forces: a pathogen’s reproductive cycle, defining its relative transmission rate upon physical proximity between individuals with full or partial susceptibility, as well as human behaviour, via the frequency of transmission-prone contacts between individuals itself [3]. Critical events such as the emergence of fitter mutants or collective shifts in human activity patterns set the pace for new epidemic waves. Real-time monitoring of these forces during an epidemic, whether it is fueled mostly by increased contact levels or changes in relative transmissibility, is of paramount value for epidemic forecasting as well as the ability to set up informed, targeted mitigation strategies and estimating the effects of (non-)pharmaceutical health policies [4].

Using SARS-CoV-2 and Influenza as key examples of airborne transmissible contagions, we showcase monitoring and forecast tools for epidemic crises centered around a crowd-sourced, real-time method to assess levels of physical proximity in a population using GPS location information, the Contact Index *CX* [5]. We show that diverging trends between contact levels and independently recorded infection surveillance are indicators of altered relative viral transmissibility. Using 2020-specific data as a baseline for purely contact-driven SARS-CoV-2 epidemics, all observed transition points are explained by the onset of key immune escape variants (alpha, delta, omicron). The resulting dual evolution, Contact Index *CX* and relative transmissibility *T*, provides a highly transparent and timely picture of ongoing epidemics, including the possibility to identify likely driving forces in future epidemic waves.

## 2. Materials and Methods

### 2.1 Contact metrics relevant for epidemics

Contact networks are a representation of human interactions [6] with immediate implications for the spread of contagions in a population [7, 8]: Nodes represent individuals and edges are drawn between pairs of nodes in the event of contact between them (Figure 1(a,b)). A contagion can propagate through a population along paths following the links of the network.

**Figure 1:**
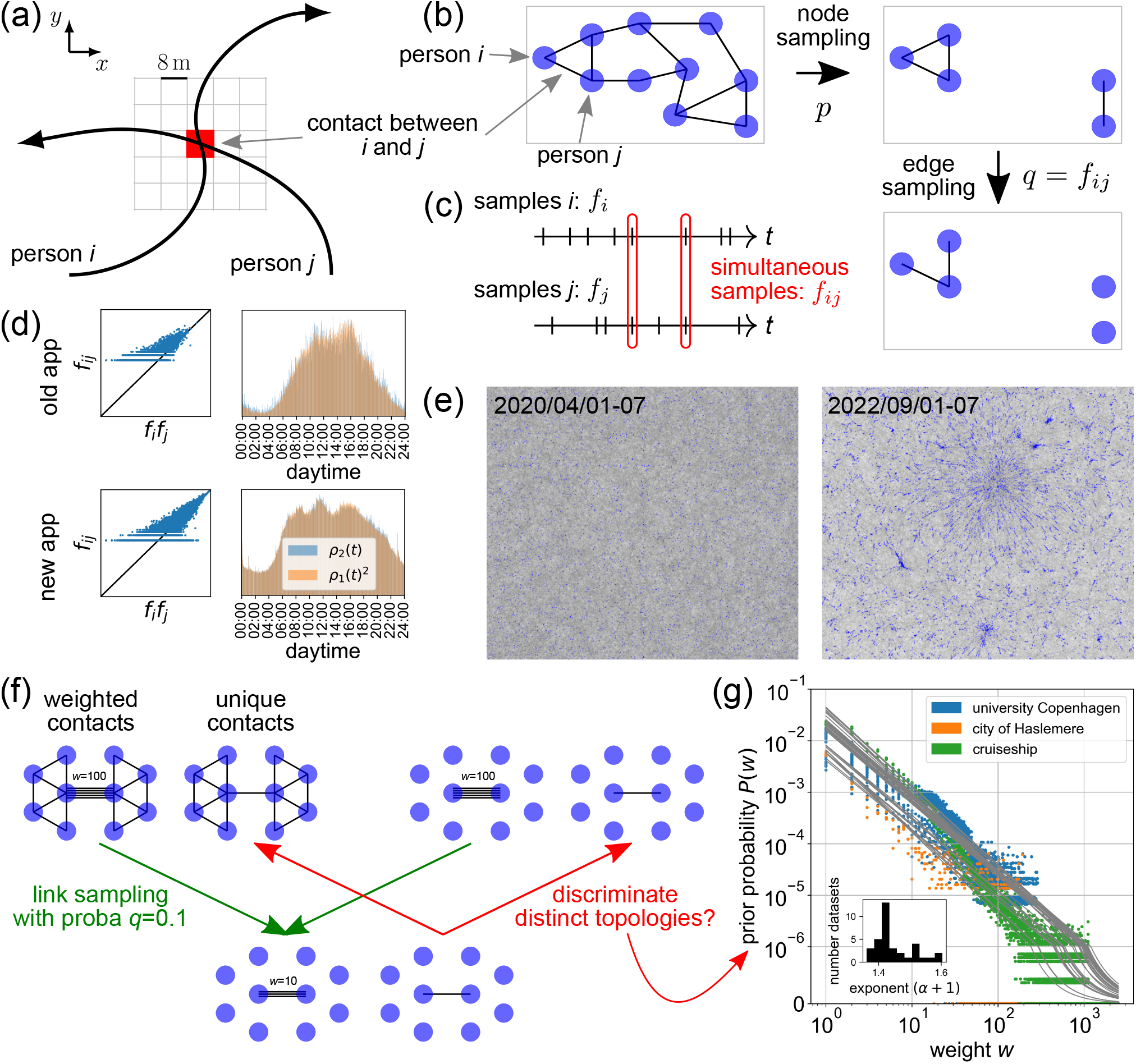
Contact networks: definition, measurement and inference. (a) Co-location of 2 (or more) devices i and j within the same 8 m ×8 m cell within 2 min defines a contact. (b) In the network, pairs of individuals/mobile devices (nodes) are connected by their contacts (edges). The network sampling induced by the data collection app retains nodes with proba p (incl. links between pairs of retained nodes), reflective of the population share of app users. Subsequently, links are retained with proba *q*. (c) Ticks along the time axes indicate samples from a pair of devices *i* and *j*. q depends on the likelihood *f*_*ij*_ of simultaneous samples (red encircled samples), a necessary condition to observe a contact between them. (d) Comparison between actual simultaneous sample rates *f*_*ij*_ and those predicted from uncorrelated single-device sample rates f_i_f_j_ (left panel) and between the distribution of simultaneous samples over the day *ρ*_2_(*t*) with the squared distribution of single-device samples *ρ*_1_(*t*)^2^ (right panel). (e) Examples of 7-day aggregated networks under lockdown (Apr 2020) and unrestricted (Sep 2022) conditions. Blue dots represent individuals, gray links the contacts. Zoom over a 2D embedding using SG-t-SNE-II [52, 53]. (f) In weighted contact networks, links are weighted by the duration/multiplicity of contact *w*_ij_ ∈ {0, 1, 2, … } between nodes *i* and *j*, while unique contact networks only distinguish between presence or absence of contact, *a*_*ij*_ ∈ {0, 1. } Example of information loss upon link sampling: Networks with distinct topologies (left vs. right set of networks) can yield similar sample networks (bottom networks) upon the same sampling process (green arrows). Discriminating distinct original networks from the sample network (red arrows) thus requires additional information. (g) Prior information is extracted from weight distributions *P* (*w*) found in complete contact networks [17, 18, 19].

Intuitively, transmission levels scale with the average number of links per node ⟨*k*⟩ = Σ_*k*≥0_ *kP*(*k*) = 2*L*/*N* [3], where *P* (*k*) is the distribution of these numbers across a network and N (L) is the number of nodes (links). Beyond this local property, more global topological network features – how contacts are collectively configured across the network – do also affect the course of epidemics [3] by fueling and constraining the number of available paths. Groundbreaking epidemiological and network-theoretical work established that the effective reproduction number *R*_eff_, quantifying epidemic spreading, scales with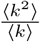i.e. the presence of very social nodes (superspreaders) with outstanding k mediate enhanced propagation. Typical social networks are very inhomogeneous in terms of social activity, with outstanding community structure and few individuals responsible for most contacts [9]. The pivotal role of the second moment ⟨*k*^2^⟩ =Σ_*k*≥0_ *k*^2^*P* (*k*) is intuited by the friendship paradox [13]: An individual’s friends are on average more social than oneself; in other words, the number of next-nearest neighbors (k^2^) in the network exceeds the expectation ⟨*k*⟩2 from the number of nearest neighbors, a mere consequence of non-zero variance in *P* (*k*): ⟨*k*^2^⟩ −⟨*k*⟩^2^ > 0 (Supp Mat S2).

### 2.2 Assessing contact levels in real-world networks

The contact network relevant to transmission of airborne viruses such as Influenza and SARS-CoV-2 arises from physical proximity between individuals (Figure 1(a)). Compared to (virtual) social networks, such real-world networks are expected to have distinct properties, as they are constrained by geography and physical distance, but are also tremendously more difficult to track at the population scale. Coarse contact and mixing patterns in real-world networks have been inferred using limited data gathered from surveys [14, 15] or viral phylogeny [16]. Locally confined real-world networks, such as on cruise ships [17], school campuses [18] or within towns [19] have been measured using Bluetooth communication between nearby mobile devices.

We use a previously developed approach to probe population-scale real-world contact networks based on crowd-sourced datasets of GPS locations [20, 5] to measure the Contact Index 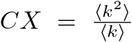 as a statistical measure of contact levels relevant for epidemics [5]. The crowd-sourcing data is collected in near real-time via opt-in from each of an anonymized panel of 1 million mobile app users (roughly 1 % of Germany’s population) and consists of ≈ 100 daily samples per device tagged with time and GPS location information. It allows us to reconstruct samples of the actual contact network realized in the population: Contacts (links) are drawn between devices (nodes) co-located in space and time (Figure 1(a) and Supp Mat S1). Examples of reconstructed contact networks are shown in Fig. 1(e).

### 2.3 Network sampling correction

The incomplete nature of such crowd-sourced data represents a major challenge: Contacts from uninvolved or inactive devices are not captured, giving rise to missing nodes and links in the network. This aspect of our data can be crafted into a network sampling framework [21, 22] in which nodes and edges are randomly removed with probabilities p and q, respectively (Figure 1(b,c) and Supp Mat S3). p denotes the population share represented in the panel of app users, while q is interpreted as the rate *f*_*ij*_ of simultaneous samples from pairs of app users (Figure 1(c)), a necessary condition to detect a contact between users with individual sample rates f_i_ and f_j_, respectively. These sampling parameters are subject to change over time beyond daytime-related periodicity (see below), mostly in response to software updates and app usage (Figure 1(d)), and are heterogeneous in space (Supp Mat S4 and Figure S3(a,b,c)).

For simplicity, we here use daily averages of sample rates. The rate *f*_*ij*_ of simultaneous samples tends to exceed the expectation from individual frequencies *f*_*i*_*f*_*j*_ under the hypothesis of independence of distinct mobile devices, i.e. *f*_*ij*_ > *f*_*i*_*f*_*j*_, especially prior to February 2020 (Figure 1(d)); a major app update in February 2020 has significantly altered the daytime distribution and overall number of samples (Figure 1(d)). This apparent correlation between devices stems from the non-uniformity of the sampling activity over the day: Devices are more active during daytime than at night, an effect particularly prominent prior to February 2020 (Figure 1(d)). However, aside from a common daytime pattern, devices show a predominantly independent activity pattern from one another (Figure 1(d)): At any given timepoint (2 min interval), squared single-device distributions, i.e.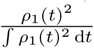do capture the distribution of simultaneous samples *ρ*_2_(*t*) across the day well. Solely in consequence to the daytime-related correlation, we are likely to slightly underestimate the true value of *q* by using daily averages.

Our improved mathematical modeling based on Horvitz-Thompson theory disentangles actual changes in contact levels from signals unrelated to the users’ contact behaviour, including participation and activity levels in the user panel, but excluding correlation between devices, see above. We thus achieve a persistent and comparable results across the full time span since the beginning of measurement in 2019 (Supp Mat S3 and Supp Mat S4). In summary, we show that the Contact Index CX of an unobserved complete network *G* can be retrieved from a network sample *G*^∗^ obtained under the described sampling scheme according to

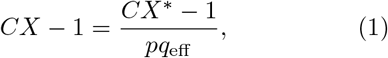

where 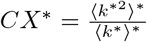 is the same quantity measured within the network sample and *q*_eff_ is an effective node sampling probability for networks of unique contacts (see below).

Importantly, abstractions of contact networks exist in two distinct flavours: weighted versus unweighted [23]. Links may be endowed with weights *w*_*ij*_ ∈ {0, 1, 2, … } representing the duration or multiplicity of contact between individuals *i* and *j* [24] or simply indicate the presence or absence of contact *a*_*ij*_ = sgn(*w*_*ij*_) ∈ {0, 1} (Figure 1(f)). In the epidemiological context, we assume that network topology, represented by *a*_*ij*_, is more important than the recurrence of contacts between the same individuals: For instance, the (statistical) contribution to viral spread from a cluster of short contacts at a crowded event would outpace a lengthy contact between an isolated couple while in lockdown. We thus focus on unweighted networks and exclude contact duration in our analyses other than in the fact that short contacts are unlikely to be recorded during the random sampling inherent to the crowd-sourcing method.

However, network sampling destroys topological information about underlying complete networks (Figure 1(f)); the success of Horvitz-Thompson theory [21] to establish a connection between original and sample networks relies in the use of weighted links (Supp Mat S3). To establish the same connection for unweighted networks, we devised a Bayesian approach which identifies missing topological information as the weight distribution for existing links in the complete network P (*w*|*w* > 0) and defines the edge sampling probability as

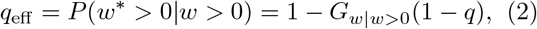

where *G*_*w*|*w*>0_(ξ) = Σ_*w*>0_ *P* (*w*|*w* > 0)ξ^*w*^ is the probability generating function of *P* (*w*|*w* > 0) (Supp Mat S3). We find that available complete real-world networks in various contexts [17, 18, 19] appear to show strikingly similar weight distributions (Figure 1(g)), which suggests a universal shape of *P* (*w*|*w* > 0) also applicable to our problem. Here, “complete” refers to the aspect that these networks represent a fraction of the population (*p* < 1), but all contacts within that sub-population are being detected (*q* = 1) – node sampling, but no edge sampling. These distributions are consistent with power laws *P* (*w*|*w* > 0) = *w*^−(1+α)^/?(1 + α) with small exponents [25, 26] (Figure 1(g)), a repeatedly demonstrated feature of complex networks [27] and beyond [28]. Yet, we do not imply that power laws are the true mechanism behind network weights, as a variety of other distribution classes are easily confounded with power laws [28, 29, 30], but merely use it as a prior for *P* (*w*|*w* > 0).

## Results

### Evolution of CX since 2019

By means of our refined correction method for network sampling effects, we achieve a consistent measurement of contact levels since the beginning of crowd-sourcing in 2019, despite the time-dependent sampling. That is, we cover the prelude and entire course of the SARS-CoV-2 epidemic in Germany (Figure 2(a)). The gap in February 2020 is explained by missing data due to the rollout of a major crowd-sourcing software update.

**Figure 2:**
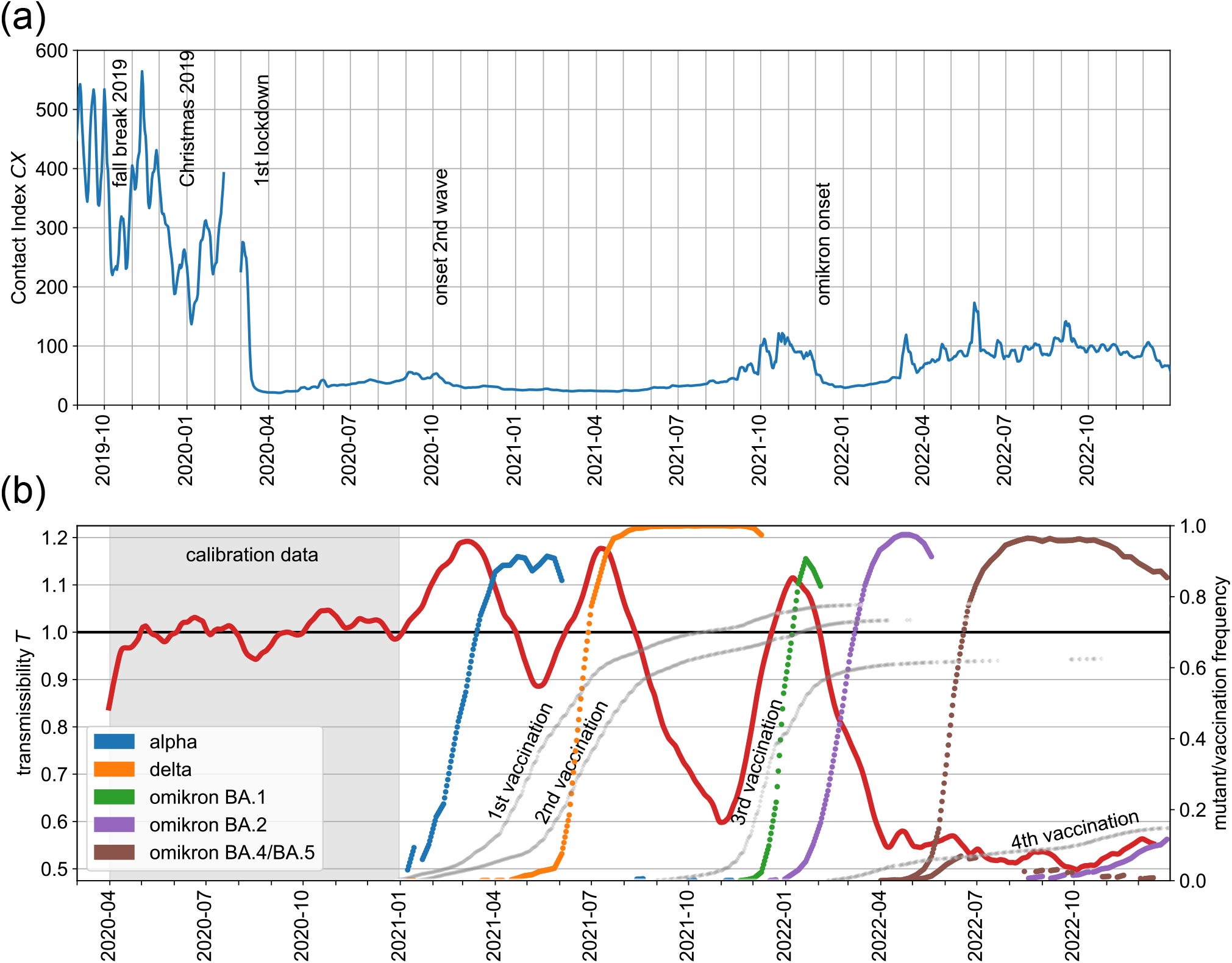
Real-time observation of driving forces in SARS-CoV-2 epidemics: contact levels and relative transmissibility. (a) Evolution of the Contact Index CX = ⟨*k*^2^⟩/ ⟨*k*⟩in Germany over the course of > 3 years (2019-2022), carrying the signature of various collective behaviour changes in response to the epidemic situation (as indicated). The gap in February 2022 is explained by a major app update. (b) The slowly varying relative transmissibility ⟨*T*⟩ (*t*) (red) quantifying the intrinsic efficiency of SARS-CoV-2 transmission, measured from the ratio of reproduction numbers (*R*_eff_) and contact levels (*CX*), see Eq. (3). The gray-shaded time interval is wild-type dominated and was used to calibrate CX from our crowd-sourcing method and *R*_eff_ from infection surveillance (Figure S1(a, inset)). The rising frequencies of key SARS-CoV-2 immune escape variants (colored lines, see legend) and well as of vaccine status in Germany (light gray lines) are shown (right axis).

Holiday season comes along with reduced *CX* under normal conditions, as shown by the Fall and Christmas breaks in 2019, thus showing a reduction of transmission-prone contacts. The onset of the first SARS-CoV-2 wave in March 2020 induced an unequivocally more pronounced drop in *CX*, probably explained by a more systematic cessation of super-spreading activities. The dramatically altered contact network structure during a lockdown is depicted in Figure 1(e).

Since onset of the SARS-CoV-2 pandemic, changes in contact behaviour as reflected by *CX* underwent several periods of spiking (partial or complete deregulation of mass events in fall 2020, fall 2021 and spring 2022) and damping (winter wave 2020, emergence of the omicron variant in late 2021). Overall, a similar evolution is observed between *CX* and the rigor of SARS-CoV-2-related policy as measured by the Government-Response Index [31] (Figure S1(a)), thus indicating broad awareness of the situation at the population and governance levels albeit no causal link shall be implied.

Interestingly, recent *CX* values have not yet returned to pre-pandemic levels by a factor of 2 to 3, despite a return to no contact-related restrictions in 2022. This suggests the existence of a hysteresis effect in addition to the fast response of *CX* discussed above: The collective behaviour has not returned to its unperturbed state in response to relaxed conditions, possibly as a result of continued broad perception of disease risk [32, 33].

From a dimensional viewpoint, *CX* represents an average number of (next-nearest) contacts per (nearest) contact: Comparing values of *CX* across areas with vastly different population densities within Germany supports our expectation that *CX* scales (non-linearly) with the absolute propensity of physical proximity between individuals (Figure S3(d) and Supp Mat S4).

### 3.2 Deciphering epidemic forces: contacts vs. relative transmissibility

In 2020, SARS-CoV-2 epidemic trends were primarily driven by trends in contact levels, as both immune escape variants and vaccines were not yet relevant and relative SARS-CoV-2 transmissibility its intrinsic transmission probability per contact was thus constant (Figure 2(b)): Official daily now-cast reproduction numbers *R*_eff_, independently recorded from national infection surveillance [34], correlate well with daily CX, but *CX* shows a time lead of approximately 2 − 3 weeks over *R*_eff_ (Figure S1(a, right inset)) [5], explained by incubation time as well as testing and reporting delays. This underlines the predictive character of real-time contact metrics for wild-type dominated epidemics [20]. Since then, the correlation between *R*_eff_ and *CX* has repeatedly changed, with the resulting signal quantifying shifts in relative transmissibility accountable to key epidemic changes other than contacts.

The effective reproduction number *R*_eff_ is defined by *R*_eff_ = ⟨*k*⟩ · *U* · τ, where ⟨*k*⟩ denotes the contact number per day, *U* the probability of transmission per contact, and τ the mean duration of infectivity in days. Both *U* and τ are determined by physiological processes involved in transmission and, together, define the intrinsic transmission efficiency (per contact) *T* = *U* · τ.

Furthermore, as we assume 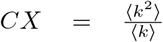 replaces ⟨*k*⟩, we replace the definition by *R*_eff_ = (a + b · *CX*) · T. A linear relationship of this form between *CX* and *R*_eff_ is motivated by our findings in 2020. We use values for a and b obtained from a linear regression between *CX* and wild-type *R*_eff_ data at the optimal time delay of ∆t = 16 days (Figure S1(a, left inset) and Supp Mat S5). Upon interpreting *R*_WT_(*CX*) ≡ a+b·*CX* as the wild-type specific reproduction number, we have that

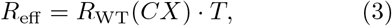

where *T* represents relative transmissibility with respect to wild-type in a fully susceptible population (*T*_WT_ = 1). Note that, in contrast to now-cast data, Eq. (3) assigns reproduction numbers to the day of contact/infection.

From independently recorded values for *R*_eff_ and *CX*, we can determine the relative transmissibility of the contagion by factoring out contact-related contributions from overall infection dynamics as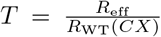for any given day. We expect network-wide propagation of transmissibility-related information to be slow compared to network dynamics itself and, thus, *T* to undergo evolution on longer timescales. We interpret fast signal in *T* as random fluctuations from the measurement of *R*_eff_ and capture actual trends by ⟨*T*⟩, centered averages over sliding time windows of 2 months (Supp Mat S5).

### 3.3 Epidemic evolution of relative SARS-CoV-2 transmissibility

The evolution of relative SARS-CoV-2 transmissibility ⟨*T*⟩ is shown in Figure 2(b). This time series reenacts the various phases of the SARS-CoV-2 pandemic:

Relative SARS-CoV-2 transmissibility ⟨T⟩ is approximately equal to unity throughout 2020, an initial period purely driven by unperturbed wildtype epidemics that we used to “calibrate” *CX* and *R*_eff_ which evolve on shorter timescales. It subsequently follows a tug-of-war pattern shaped by alternating epidemic forces beyond contacts: immune escape variants and development of population immunity through infection and vaccination. Three waves of increased relative transmissibility are explained by the takeover of fitter virus lineages (Figure 2(b)), specifically alpha (spring 2021), delta (summer 2021) and omicron BA.1/BA.2 (winter 2021/22). We hypothesize that subsequent relaxation of ⟨*T*⟩ after each wave may be attributed to natural immunity, while the superposed long-term downward trend may be explained by the additional immunity acquisition through (initial and booster) vaccination campaigns. Interestingly, the effect of omicron BA.4/BA.5 takeover in summer 2022 on ⟨*T*⟩ is nowhere close to those of previous variants.

Comparing correlations with different parameters rules out the possibility that the measured ⟨*T*⟩ is shaped by factors confounding the reproduction numbers or *CX* values (Figure S1(b,c) and Supp Mat S5). These possible confounders include viral prevalence, *CX* itself through higher-order effects from network sampling not captured by our modeling and other topological network features (such as clustering, small-world properties) as well as *R*_eff_ itself through changes in testing strategies and systematic under-reporting of infections [35]. For instance, testing individuals indiscriminately versus focusing test capacities on suspected infection cases may lead to incomparable snapshots of ongoing infection dynamics. Overall, strong positive correlation is exclusively observed between ⟨*T*⟩ and variant dynamics (Figure S1(b,c)) [36]. In this analysis, we use test positivity [37] and results from local prevalence studies [38] as proxies for overall prevalence. Also, we neglect possible effects from network sampling on different topological measures [39, 40], but we expect trends to be conserved as long as the sampling process remains unchanged.

We note the absence of seasonal oscillations in ⟨*T*⟩ as well as clear signatures of mask mandates (in effect across many social contexts between April 2020 and April 2022). A seasonal oscillation in ⟨*T*⟩, larger values in winter and smaller values in summer, might be expected from the shift of human activity between in- and outdoor settings. Also, previous research established the effectiveness of mask usage at reducing transmission of respiratory diseases (reviewed in [41]). Overall, our results suggest that, at least in the epidemic stage of SARS-CoV-2, infection rates were predominantly driven by the strong variability in contacts as well as the repeated emergence of more transmissible variants, in line with previous findings [42, 43, 44].

#### 3.4 Forecast of infection level and trend changes

The challenge of epidemic forecast consists in the accurate prediction of current and future reproduction numbers *R*_eff_. Using the rationale that trends in infection levels carry the combined signature of trends in contact and relative transmissibility levels, we propose to construct predictions according to

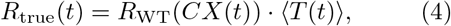

where *R*_true_ is assigned to the projected day of contact/infection. The key difference to Eq. (3) is the use of ⟨T⟩which eliminates noise from reproduction numbers. Importantly, we therefore expect that our prediction *R*_true_ represents actual epidemic trends (ground truth) more accurately than epidemic surveillance (*R*_eff_).

Figure 3(a) shows *R*_true_ together with data from infection surveillance, both plotted with respect to their date of recording (assuming real-time *CX* measurement). This shows how our prediction overall anticipates current epidemic trends that are observed via infection surveillance only about ∆*t* = 2 − 3 weeks later. Thus, we propose to use our method as a tool for real-time infection surveillance.

**Figure 3:**
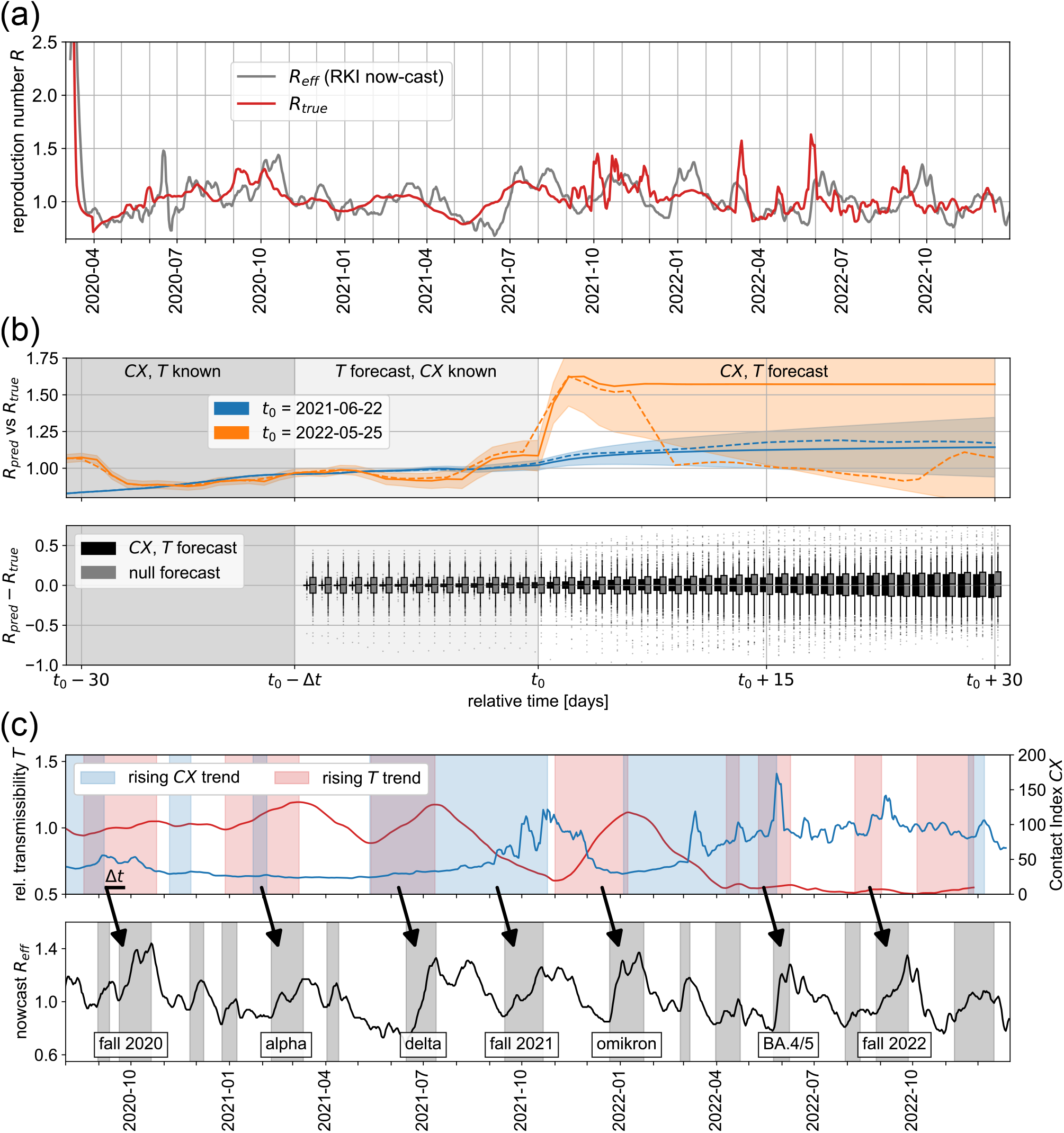
Forecast of reproduction numbers and trends from contact and transmissibility levels. (a) Comparison of SARS-CoV-2 effective reproduction numbers *R*_eff_ from infection surveillance (gray) and projected *R*_true_ using Eq. (4) (red). All reproduction numbers are assigned to their day of recording. (b, upper panel) Forecast R_pred_ of current and future SARS-CoV-2 reproduction numbers and their uncertainties (solid lines and shaded bands, respectively) using Eq. (4) and the *CX* and ⟨*T*⟩ time series. Comparison with actual *R*_true_ values (dashed lines). Denoting the current day by t_0_, *R*_eff_ and ⟨*T*⟩ are available up to t_0_ − ∆t, while *CX* is near real-time (available up to t_0_); the time series are projected beyond their last time points using ARIMA models. The forecast is shown for different choices of the current day t_0_ (see legend). (b, lower panel) The distribution of residuals between forecasted R_pred_ and actual *R*_true_ values over all choices of t_0_ over the course of 2 years (black box plots). Comparison to residuals from null projections of *R*_eff_ that make no use of *CX* (gray box plots), i.e. simple ARIMA model-based projection of infection surveillance data. The boxes indicate quartiles, while whiskers cover 90 % of the data. (c) Identification of rising trends in both contact levels and transmission efficiency (upper panel) and their relation to rising trends in *R*_eff_ (lower panel).

To extend forecasts beyond this horizon and predict future reproduction numbers, *CX* and ⟨*T*⟩ themselves need to be projected beyond latest data.

For several choices of the current day t_0_, Figure 3(b) showcases forecasts (*R*_pred_) where *CX* and ⟨*T*⟩ are continued beyond the last days of available data (*t*_0_ and *t*_0_ − ∆*t*, respectively) using autoregressive integrated moving average (ARIMA) models prior to applying Eq. (4) (Supp Mat S6). These forecasts outperform a null forecast based on a mere ARIMA-type continuation of infection surveillance data (*R*_eff_), as shown by narrower distributions of residuals (*R*_pred_−*R*_true_) across all choices of t_0_ (Figure 3(b)). Furthermore, we highlight the broad applicability of our method to airborne infectious diseases by performing an identical forecast analysis for Influenza (Figure S2(a)), using coarser infection surveillance data [45] and presuming a similar relationship between *R*_eff_ and *CX* as for SARS-CoV-2 (Supp Mat S6).

Most importantly, trend changes in epidemic driving forces such as ⟨*T*⟩ and *CX* are indicators of new phases in an epidemic. Timely detection of new trends in these time series, e.g. using anomaly detection methods, can provide valuable information to estimate the risk of upcoming epidemic waves and to predict their nature – whether dynamics is fueled by contacts or increased transmission efficiency. Such trend detection is potentially easier to achieve but equally informative than the ability to accurately predict infection surveillance. The onset of rising trends could shape decision-making with regard to the effectiveness of health policies, e.g. pharmaceutical and non-pharmaceutical interventions for rising ⟨*T*⟩ and *CX*, respectively. Figures 3(c) and S2(b) highlight rising and falling trends in both *CX* and T for SARS-CoV-2 and Influenza, respectively, akin to trends in stock prices. For SARS-CoV-2, trend changes are timely indicators of all major escape variant- and contact-driven epidemic turning points (Figure 3(c)). Unlike for SARS-CoV-2 in its epidemic stage, major upheavals in relative transmissibility for Influenza are limited to seasonality, with the notable exception of 2020, presumably reflecting its endemic dynamics (Figure S2(b)).

## Discussion

We presented a simple, yet insightful quantitative method for a data-driven decomposition of overall epidemic dynamics into contact-related and transmission efficiency-related contributions. It relies on both the availability of infection surveillance data as well as crowd-sourced GPS location data to detect and quantify physical proximity between susceptible individuals. Its appeal resides in the merely bivariate yet highly informative projection of epidemics paving the way towards timely identification of driving forces in an ongoing epidemic human versus viral factors – and possibly effective mitigation strategies – pharmaceutical versus non-pharmaceutical.

The approach can be used for epidemic forecast in multiple ways. Recent and projected future values of *CX* and ⟨*T*⟩ can be used for short-term (2 − 3 weeks) and long-term prediction of infection or reproduction numbers, thus taking our previously described short-term forecast further [5]. Yet, a timely detection of trend changes could reliably forecast upcoming waves and their nature without the necessity to accurately predict infection surveillance data. These tools can lead towards a more strategic approach to epidemic mitigation and potentially save lives by reducing the spread of deadly diseases.

Results from the presumably most systematically tracked epidemic to date, SARS-CoV-2, draw the picture of co-evolution within the virus-host relation: Increasing immunity levels in the host population alternate with step-wise adaptation of the virus through immune-escape variants. Other frequently discussed factors, including mask policies and seasonality, are presumably still below the current statistical resolution of our method, defined by the sampling noise in the *CX* and *R*_eff_ time series. Moreover, a larger impact of seasonal variation is expected in the endemic phase of SARS-CoV-2 [46].

Our method is broadly applicable to airborne contagions beyond SARS-CoV-2, but depends on the availability of infection surveillance and crowdsourcing strategies that remain persistent over extended amounts of time. Changes in testing strategy can lead to signal and biases unrelated to underlying epidemic driving forces [35]. More crucially, systematic infection surveillance is not implemented beyond the case of SARS-CoV-2. We illustrated a framework to correct for the effect of varying sampling depth in the contact network. Yet, higher-order effects in the signal can occur as a result of sampling aspects not captured by our mathematical modeling. In order to ensure valid prognoses through our method, we advocate for systematic and persistent crowd-sourcing and infection surveillance strategies across a variety of diseases with epidemic potential.

Geographical resolution of our forecast method is currently limited by the sampling depth, as the estimation especially of higher moments of degree distributions P ⟨*k*⟩ becomes increasingly difficult as smaller portions of the network are available. A higher spatial resolution of contact and relative transmissibility levels, with potential to locate the origin of new variants of concern and define locally targeted mitigation strategies, can be achieved by e.g. increasing the panel of app users.

Our analysis assumes statics, but actual contact networks are dynamic in nature [47, 48]: While some contacts are frequently repeated (e.g. between household members), other contacts are randomly redrawn on each occasion (e.g. in public transportation), with implications for epidemic spread [49, 50]. Our method can be improved by analyzing contact data in light of existing models of dynamic networks [51, 48].

## Data Availability

All aggregated data produced in the present study (i.e., anonymized daily contact networks, daily participant and sample numbers) are available upon reasonable request to the authors. Official infection surveillance, vaccine coverage, and virus variant data for Germany used in this work are publicly available from the Robert Koch Institute (see references in the manuscript).

## Acknowledgment

This work was supported by grants from the Federal Government of Germany through the Federal Ministry for Economic Affairs and Climate Action (BMWK) for the project DAKI-FWS (01MK21009A) and the Federal Ministry of Education and Research (BMBF) for the project Optim-Agent (031L0299).

## References

[1] A. Rodríguez, H. Kamarthi, P. Agarwal, J. Ho, M. Patel, S. Sapre, B. A. Prakash, Data-centric epidemic forecasting: A survey, arXiv preprint arXiv:2207.09370 (2022).

[2] A. Maxmen, Has covid taught us anything about pandemic preparedness?, Nature 596 (2021) 332–335.

[3] R. Pastor-Satorras, C. Castellano, P. Van Mieghem, Vespignani, Epidemic processes in complex networks, Rev. Mod. Phys. 87 (2015) 925–979. doi:10.1103/RevModPhys.87.925. URL https://link.aps.org/doi/10.1103/RevModPhys.87.925

[4] T. Alamo, D. G. Reina, P. Millén Gata, V. M. Preciado, G. Giordano, Data-driven methods for present and future pandemics: Monitoring, modelling and managing, Annual Reviews in Control 52 (2021) 448–464. doi: https://doi.org/10.1016/j.arcontrol.2021.05.003. URL https://www.sciencedirect.com/science/article/pii/S1367578821000419

[5] S. Rädiger, S. Konigorski, A. Rakowski, J. A. Edelman, D. Zernick, A. Thieme, C. Lippert, Predicting the sars-cov-2 effective reproduction number using bulk contact data from mobile phones, Proceedings of the National Academy of Sciences 118 (31) (2021) e2026731118. arXiv: https://www.pnas.org/doi/pdf/10.1073/pnas.2026731118, doi:10.1073/pnas.2026731118. URL https://www.pnas.org/doi/abs/10.1073/pnas.2026731118

[6] M. E. J. Newman, J. Park, Why social networks are different from other types of networks, Phys. Rev. E 68 (2003) 036122. doi:10.1103/PhysRevE.68.036122. URL https://link.aps.org/doi/10.1103/PhysRevE.68.036122

[7] M. Keeling, The implications of network structure for epidemic dynamics, Theoretical population biology 67 (1) (2005) 1–8.

[8] C. Moore, M. E. J. Newman, Epidemics and percolation in small-world networks, Phys. Rev. E 61 (2000) 5678–5682. doi:10.1103/PhysRevE.61.5678. URL https://link.aps.org/doi/10.1103/PhysRevE.61.5678

[9] M. Barthélemy, A. Barrat, R. Pastor-Satorras, A. Vespignani, Dynamical patterns of epidemic outbreaks in complex heterogeneous networks, Journal of Theoretical Biology 235 (2) (2005) 275–288. doi:https://doi.org/10.1016/j.jtbi.2005.01.011. URL https://www.sciencedirect.com/science/article/pii/S0022519305000251

[10] Y. Moreno, R. Pastor-Satorras, A. Vespignani, Epidemic outbreaks in complex heterogeneous networks, The European Physical Journal B-Condensed Matter and Complex Systems 26 (4) (2002) 521–529.

[11] R. M. May, R. M. Anderson, M. E. Irwin, R. M. Anderson, J. M. Thresh, The transmission dynamics of human immunodeficiency virus (hiv), Philosophical Transactions of the Royal Society of London. B, Biological Sciences 321 (1207) (1988) 565–607. arXiv: https://royalsocietypublishing.org/doi/pdf/10.1098/rstb.1988.0108, doi:10.1098/rstb.1988.0108. URL https://royalsocietypublishing.org/doi/abs/10.1098/rstb.1988.0108

[12] R. M. May, R. M. Anderson, Transmission dynamics of hiv infection, Nature 326 (1987) 137–142.

[13] S. L. Feld, Why your friends have more friends than you do, American Journal of Sociology 96 (6) (1991) 1464–1477. arXiv:https://doi.org/10.1086/229693, doi:10.1086/229693. URL https://doi.org/10.1086/229693

[14] J. M. Read, K. T. Eames, W. J. Edmunds, Dynamic social networks and the implications for the spread of infectious disease, Journal of The Royal Society Interface 5 (26) (2008) 1001–1007.

[15] J. Mossong, N. Hens, M. Jit, P. Beutels, K. Auranen, R. Mikolajczyk, M. Massari, S. Salmaso, G. S. Tomba, J. Wallinga, J. Heijne, M. Sadkowska-Todys, M. Rosinska, W. J. Edmunds, Social contacts and mixing patterns relevant to the spread of infectious diseases, PLOS Medicine 5 (3) (2008) 1–1. doi:10.1371/journal.pmed.0050074. URL https://doi.org/10.1371/journal.pmed.0050074

[16] G. E. Leventhal, R. Kouyos, T. Stadler, V. von Wyl, S. Yerly, J. Böni, C. Cellerai, T. Klimkait, H. F. Gänthard, S. Bonhoeffer, Inferring epidemic contact structure from phylogenetic trees, PLOS Computational Biology 8 (3) (2012) 1–10. doi:10.1371/journal.pcbi.1002413. URL https://doi.org/10.1371/journal.pcbi.1002413

[17] R. Pung, J. A. Firth, L. G. Spurgin, V. J. Lee, A. J. Kucharski, Using high-resolution contact networks to evaluate sars-cov-2 transmission and control in large-scale multi-day events, Nature communications 13 (1) (2022) 1–11.

[18] P. Sapiezynski, A. Stopczynski, D. D. Lassen, S. Lehmann, Interaction data from the copenhagen networks study, Scientific Data 6 (1) (2019) 315.

[19] S. M. Kissler, P. Klepac, M. Tang, A. J. Conlan, J. R. Gog, Sparking “the bbc four pandemic”: Leveraging citizen science and mobile phones to model the spread of disease, bioRxiv (2020). arXiv:https://www.biorxiv.org/content/early/2020/05/12/479154.full.pdf, doi:10.1101/479154. URL https://www.biorxiv.org/content/early/2020/05/12/479154

[20] F. W. Crawford, S. A. Jones, M. Cartter, S. G. Dean, J. L. Warren, Z. R. Li, J. Barbieri, J. Campbell, P. Kenney, T. Valleau, O. Morozova, Impact of close interpersonal contact on covid-19 incidence: Evidence from 1 year of mobile device data, Science Advances 8 (1) (2022) eabi5499. arXiv:https://www.science.org/doi/pdf/10.1126/sciadv.abi5499, doi:10.1126/sciadv.abi5499. URL https://www.science.org/doi/abs/10.1126/sciadv.abi5499

[21] E. D. Kolaczyk, Statistical Analysis of Network Data, Springer New York, NY, 2009. doi:https://doi.org/10.1007/978-0-387-88146-1.

[22] P. Hu, W. C. Lau, A survey and taxonomy of graph sampling, CoRR abs/1308.5865 (2013). arXiv:1308.5865. URL http://arxiv.org/abs/1308.5865

[23] M. A. Serrano, M. Boguñá, R. Pastor-Satorras, Correlations in weighted networks, Phys. Rev. E 74 (2006) 055101. doi:10.1103/PhysRevE.74.055101. URL https://link.aps.org/doi/10.1103/PhysRevE.74.055101

[24] A. Barrat, M. Barthélemy, R. Pastor-Satorras, A. Vespignani, The architecture of complex weighted networks, Proceedings of the National Academy of Sciences 101 (11) (2004) 3747–3752. arXiv:https://www.pnas.org/doi/pdf/10.1073/pnas.0400087101, doi:10.1073/pnas.0400087101. URL https://www.pnas.org/doi/abs/10.1073/pnas.0400087101

[25] H. Ebel, L.-I. Mielsch, S. Bornholdt, Scale-free topology of e-mail networks, Phys. Rev. E 66 (2002) 035103. doi: 10.1103/PhysRevE.66.035103. URL https://link.aps.org/doi/10.1103/PhysRevE.66.035103

[26] M. E. J. Newman, Scientific collaboration networks. i. network construction and fundamental results, Phys. Rev. E 64 (2001) 016131. doi:10.1103/PhysRevE.64.016131. URL https://link.aps.org/doi/10.1103/PhysRevE.64.016131

[27] R. Guimera, S. Mossa, A. Turtschi, L. N. Amaral, The worldwide air transportation network: Anomalous centrality, community structure, and cities’ global roles, Proceedings of the National Academy of Sciences 102 (22) (2005) 7794–7799.

[28] A. Clauset, C. R. Shalizi, M. E. J. Newman, Power-law distributions in empirical data, SIAM Review 51 (4) (2009) 661–703. arXiv:https://doi.org/10.1137/070710111, doi:10.1137/070710111. URL https://doi.org/10.1137/070710111

[29] J.-D. J. Han, D. Dupuy, N. Bertin, M. E. Cusick, M. Vidal, Effect of sampling on topology predictions of protein-protein interaction networks, Nature biotechnology 23 (7) (2005) 839–844.

[30] R. Perline, Strong, weak and false inverse power laws, Statistical Science 20 (1) (2005) 68–88. URL http://www.jstor.org/stable/20061161

[31] T. Hale, N. Angrist, R. Goldszmidt, B. Kira, A. Petherick, T. Phillips, S. Webster, E. Cameron-Blake, L. Hallas, S. Majumdar, et al., A global panel database of pandemic policies (oxford covid-19 government response tracker), Nature human behaviour 5 (4) (2021) 529–538.

[32] C. Betsch, S. Eitze, P. Sprengholz, L. Korn, P. Shamsrizi, M. Geiger, E. Sievert, L. Lehrer, M. Jenny, Zusammenfassung und empfehlungen welle 69 (2022). URL https://projekte.uni-erfurt.de/cosmo2020/web/summary/69/

[33] C. Betsch, L. Wieler, M. Bosnjak, M. Ramharter, V. Stollorz, S. Omer, L. Korn, P. Sprengholz, L. Felgendreff, S. Eitze, P. Schmid, Germany covid-19 snap-shot monitoring (cosmo germany): Monitoring knowledge, risk perceptions, preventive behaviours, and public trust in the current coronavirus outbreak in germany (Mar. 2020). doi:10.23668/psycharchives.2776. URL https://psycharchives.org/index.php/en/item/e5acdc65-77e9-4fd4-9cd2-bf6aa2dd5eba

[34] M. an der Heiden, Sars-cov-2-nowcasting und -r-schaetzung (Jan. 2023). doi:10.5281/zenodo.7571376. URL https://doi.org/10.5281/zenodo.7571376

[35] H. Rossman, E. Segal, Nowcasting the spread of sars-cov-2, Nature microbiology 7 (1) (2022) 16–17.

[36] R. Koch-Institut, Anzahl und anteile von voc und voi in deutschland (2023). URL https://www.rki.de/DE/Content/InfAZ/N/Neuartiges_Coronavirus/Daten/VOC_VOI_Tabelle.xlsx?blob=publicationFile

[37] E. Mathieu, H. Ritchie, L. Rodés-Guirao, C. Appel, C. Giattino, J. Hasell, B. Macdonald, S. Dattani, D. Beltekian, E. Ortiz-Ospina, M. Roser, Coronavirus pandemic (covid-19) (2020). URL https://ourworldindata.org/coronavirus

[38] H. Neuhauser, N. Buttmann-Schweiger, J. Fiebig, C. Poethko-Mäller, F. Prätz, G. Sarganas Margolis, R. Thamm, M. Zimmermann, Observatorium serologischer Studien zu SARS-CoV-2 in Deutschland (Sep. 2022). doi:10.5281/zenodo.7043025. URL https://doi.org/10.5281/zenodo.7043025

[39] J. D. Noh, Percolation transition in networks with degree-degree correlation, Phys. Rev. E 76 (2007) 026116. doi:10.1103/PhysRevE.76.026116. URL https://link.aps.org/doi/10.1103/PhysRevE.76.026116

[40] S. H. Lee, P.-J. Kim, H. Jeong, Statistical properties of sampled networks, Phys. Rev. E 73 (2006) 016102. doi:10.1103/PhysRevE.73.016102. URL https://link.aps.org/doi/10.1103/PhysRevE.73.016102

[41] J. T. Brooks, J. C. Butler, Effectiveness of Mask Wearing to Control Community Spread of SARS-CoV-2, JAMA 325 (10) (2021) 998–999. arXiv: https://jamanetwork.com/journals/jama/articlepdf/2776536/jama\_brooks\_2021\_it\_210006\_1631033869.97869.pdf, doi:10.1001/jama.2021.1505. URL https://doi.org/10.1001/jama.2021.1505

[42] F. Balloux, C. Tan, L. Swadling, D. Richard, C. Jenner, M. Maini, L. van Dorp, The past, current and future epidemiological dynamic of SARS-CoV-2, Oxford Open Immunology 3 (1) (06 2022). arXiv:https://academic.oup.com/ooim/article-pdf/3/1/iqac003/48744431/iqac003.pdf, doi:10.1093/oxfimm/iqac003. URL https://doi.org/10.1093/oxfimm/iqac003

[43] G. P. Guy Jr, F. C. Lee, G. Sunshine, R. McCord, M. Howard-Williams, L. Kompaniyets, C. Dunphy, M. Gakh, R. Weber, E. Sauber-Schatz, et al., Association of state-issued mask mandates and allowing on-premises restaurant dining with county-level covid-19 case and death growth rates—united states, march 1– december 31, 2020, Morbidity and Mortality Weekly Report 70 (10) (2021) 350.

[44] D. K. Chu, E. A. Akl, S. Duda, K. Solo, S. Yaacoub, H. J. Schänemann, D. K. Chu, E. A. Akl, A. El-harakeh, A. Bognanni, T. Lotfi, M. Loeb, A. Hajizadeh, A. Bak, A. Izcovich, C. A. Cuello-Garcia, C. Chen, D. J. Harris, E. Borowiack, F. Chamseddine, F. Schänemann, G. P. Morgano, G. E. U. Muti Schänemann, G. Chen, H. Zhao, I. Neumann, J. Chan, J. Khabsa, L. Hneiny, L. Harrison, M. Smith, N. Rizk, P. Giorgi Rossi, P. Abi-Hanna, R. El-khoury, R. Stalteri, T. Baldeh, T. Piggott, Y. Zhang, Z. Saad, A. Khamis, M. Reinap, S. Duda, K. Solo, S. Yaacoub, H. J. Schänemann, Physical distancing, face masks, and eye protection to prevent person-to-person transmission of sars-cov-2 and covid19: a systematic review and meta-analysis, The Lancet 395 (10242) (2020) 1973–1987. doi:https://doi.org/10.1016/S0140-6736(20)31142-9. URL https://www.sciencedirect.com/science/article/pii/S0140673620311429

[45] R. Koch-Institut, Survstat@rki 2.0 (2023). URL https://survstat.rki.de/

[46] J. P. Townsend, A. D. Lamb, H. B. Hassler, P. Sah, A. A. Nishio, C. Nguyen, A. D. Tew, A. P. Galvani, A. Dornburg, Projecting the seasonality of endemic covid-19, medRxiv (2022). arXiv: https://www.medrxiv.org/content/early/2022/10/07/2022.01.26.22269905.full.pdf, doi:10.1101/2022.01.26.22269905. URL https://www.medrxiv.org/content/early/2022/10/07/2022.01.26.22269905

[47] V. Sekara, A. Stopczynski, S. Lehmann, Fundamental structures of dynamic social networks, Proceedings of the National Academy of Sciences 113 (36) (2016) 9977–9982. arXiv: https://www.pnas.org/doi/pdf/10.1073/pnas.1602803113, doi:10.1073/pnas.1602803113. URL https://www.pnas.org/doi/abs/10.1073/pnas.1602803113

[48] P. Holme, J. Saramäki, Temporal networks, Physics Reports 519 (3) (2012) 97–125, temporal Networks. doi: https://doi.org/10.1016/j.physrep.2012.03.001. URL https://www.sciencedirect.com/science/article/pii/S0370157312000841

[49] J. Enright, R. R. Kao, Epidemics on dynamic networks, Epidemics 24 (2018) 88–97. doi:https://doi.org/10.1016/j.epidem.2018.04.003. URL https://www.sciencedirect.com/science/article/pii/S1755436518300173

[50] E. Valdano, L. Ferreri, C. Poletto, V. Colizza, Analytical computation of the epidemic threshold on temporal networks, Phys. Rev. X 5 (2015) 021005. doi:10.1103/PhysRevX.5.021005. URL https://link.aps.org/doi/10.1103/PhysRevX.5.021005

[51] X. Zhang, C. Moore, M. E. Newman, Random graph models for dynamic networks, The European Physical Journal B 90 (10) (2017) 1–14.

[52] N. Pitsianis, D. Floros, A.-S. Iliopoulos, X. Sun, Sg-t-sne-π: Swift neighbor embedding of sparse stochastic graphs, Journal of Open Source Software 4 (39) (2019) 1577.

[53] N. Pitsianis, A.-S. Iliopoulos, D. Floros, X. Sun, Space-land embedding of sparse stochastic graphs, in: 2019 IEEE High Performance Extreme Computing Conference (HPEC), 2019, pp. 1–8. doi:10.1109/HPEC.2019.8916505.

