## Supplementary Material for "Real-time Dissection and Forecast of Infection Dynamics during a Pandemic"

---

*2023-07-25*

### S1. Definition of contact networks and reconstruction from GPS crowd-sourcing data

To achieve a computationally feasible definition and identification of contacts, we divide space-time into tiles of $8\,\mathrm{m} \times 8\,\mathrm{m}$ and $2\,\mathrm{min}$ in size. In other words, the geographical area of interest (here: map of Germany) is binned into 8 m-by-8 m tiles and the time axis is binned into contiguous intervals of 2 min. We define a contact event as the co-location of two or more devices within the same tile, i.e. when $\geq 2$ individuals are close in space at about the same time such that airborne transmission between them is possible (Figure 1(a)). We infer co-location events in Germany using crowd-sourced GPS location information from a panel of approximately $10^6$ app users, each of which contributing on average 100 daily samples, thus giving rise to a daily raw dataset of the order of $10^8$ samples and 300 GB in size. The choice of 2 min is motivated from the expectation that exhaled, infectious droplets can linger in the air for substantial time after passage of an infectious individual.

The use of co-location within two-dimensional grid cells of $8\,\mathrm{m} \times 8\,\mathrm{m}$ excludes actual contacts between nearby individuals that are separated by cell boundaries. However, we think this represents an additional benefit to our approach while simplifying the definition of contacts: This exclusion effect induces a kernel with higher likelihood for a relevant contact on short-distanced pairs of individuals, as they are more likely to fall within the same grid cell. This meets the expectation that transmission probability is also a continuously decreasing function of distance.

We define the contact network as follows: Individuals/devices are implanted as nodes of a network and edges are drawn between any pair of individuals/devices whenever these devices are found to be in contact with each other, see Figure 1(b). To obtain a day-specific contact network, we aggregate all contact events observed on that given day to build such a contact network. The mechanism of drawing edges in the contact network can take on two flavours: 1) A new edge is drawn for each, even repeated pair of individuals. Here, the number of edges between any pair $i$ and $j$ or, equivalently, the edge weight $w_{ij} \in \{0, 1, 2, \dots\}$ between them represents the number of recurrences or the total duration of contact. We refer to such contact networks as *weighted* contact networks (Figure 1(f)). 2) A single, unweighted edge is drawn between any pair of devices found to be in contact, regardless of the number of recurrences within a day, to obtain *unique* contact networks. At most one link is possible between any pair of devices and weights $a_{ij}$ can only take on $a_{ij} \in \{0, 1\}$. Overall, we here exclude "self-contacts", i.e. $w_{ii} = a_{ii} = 0$ for all nodes $i$.

In the context of epidemics, we think that the number of unique contacts is most relevant: For instance, given $w$ contacts for a particular person, it matters whether a contact with a fixed partner is repeated $w$ times or whether these stem from single contacts with $w$ distinct contact partners. The latter situation arises to higher transmission potential. In other words, rather than the number of (non-unique) contacts in the network, it matters how these are distributed across the population and define its network topology. For the computation of the Contact Index $CX = \frac{\langle k^2 \rangle}{\langle k \rangle}$, we therefore focus on unique contact numbers $k_i = \sum_j a_{ij}$ where $a_{ij} \in \{0, 1\}$. However, unique contact networks and their scaling upon network sampling is mathematically more intricate, as portrayed in Section S3.

Finally, we assume contact networks to be undirected, i.e. that infection is equally likely to occur in both directions between any pair of devices in contact. We thus neglect potential asymmetries of network links arising from the temporal order of passage of individuals at a location, as a first individual passing can result in the infection of a second, subsequent individual passing, but not vice versa by a causality argument. Since we only allow for temporal delays of up to 2 min, we expect these asymmetries to be neglectable.

Figure 1(e) shows extracts of 2D embeddings of reconstructed 7-day aggregated unique contact network samples using the SG-t-SNE-Π [1, 2] method with the following parameter values: number of iterations: 1000, number of early exageration iterations: 250, exaggeration strength: 12, learning rate: 200 and SG-t-SNE scaling factor: 10. The comparison between April 2020 and September 2022 highlights the restructuring of contact networks achieved through adapted contact behavior.

### S2. Proof of the Contact Index

We show that the expected number of next-nearest neighbors of nodes in the contact network equals $\langle k^2 \rangle$, thus exceeding $\langle k \rangle^2$ whenever the nodes do not all have identical degree, which entails the *friendship paradox.*

We assume that contact networks are built according to the configuration model in which each node $i$ is assigned a degree $k_i$ with identical probability $P(k_i) \equiv P(k)$, independently from the degree of other nodes, thus conferring $k_i$ "stubs" to node $i$. To define the identity of connection pairs, the "stubs" are then randomly connected across nodes.

Now, the probability that a given connection from a reference node $i$ will be connected to a node of degree $k_j$, $P(k_j)$, does *not* equal $P(k)$, but is actually weighed by the number of "stubs" of the potential contact partner to reflect the higher propensity of social nodes (high $k_j$) to take up a randomly established connection,

$$P(k_j = n) = \frac{nP(k = n)}{\sum_{m \geq 0} mP(k = m)} = \frac{nP(k = n)}{\langle k \rangle}, \tag{S1}$$

where $\langle k \rangle$ is the mean degree per node in the network.

We introduce the probability generating function (PGF) $G_X(\xi) = \sum_{n \geq 0} P(X = n)\,\xi^n$ for a random variable $X$ and its well-known properties $G'_X(1) = \langle X \rangle$, $G''_X(1) = \langle X^2 \rangle - \langle X \rangle$, as well as $G_X(\xi) = G_Y(G_Z(\xi))$ when $X = \sum_{n=1}^{Y} Z_n$ where $Y$ and $Z_n$ are independently random and the $Z_n$ are identically and independently distributed.

Denoting $q = \sum_{j=1}^{k_i} k_j$ the number of next-nearest neighbors of a reference node $i$, we are thus interested in evaluating $\langle q \rangle = G'_q(1) = G'_{k_i}(G_{k_j}(1))\,G'_{k_j}(1)$ using the aforementioned identities of PGFs. We have

$$G_{k_j}(\xi) = \sum_{n \geq 0} \frac{nP(k = n)\,\xi^n}{\langle k \rangle} = \frac{\xi}{\langle k \rangle} \frac{\partial}{\partial \xi} \sum_{n \geq 0} P(k = n)\,\xi^n = \frac{\xi G'_k(\xi)}{G'_k(1)} \tag{S2}$$

and therefore $\langle q \rangle = G'_k\left(\frac{\xi G'_k(\xi)}{G'_k(1)}\right) \frac{G'_k(\xi) + \xi G''_k(\xi)}{G'_k(1)}\Big|_{\xi=1} = G'_k(1) + G''_k(1) = \langle k^2 \rangle$, which concludes the proof.

### S3. Estimation of $CX$ from contact network samples

#### *S3.1. Problem statement*

The Contact Index $CX = \frac{\langle k^2 \rangle}{\langle k \rangle}$, introduced and motivated in the main text, requires estimating the first two moments of the distribution of unique contact numbers in the contact network $G$ with $N$ nodes

$$\langle k \rangle = \frac{1}{N} \sum_{i \in G} k_i = \frac{1}{N} \sum_{ij \in G} a_{ij},$$

$$\langle k^2 \rangle = \frac{1}{N} \sum_{i \in G} k_i^2 = \frac{1}{N} \sum_{ij\ell \in G} a_{ij} a_{i\ell}, \tag{S3}$$

where $k_i = \sum_{j \in G} a_{ij}$ is the unique contact number (degree) of node $i$ and $a_{ij} \in \{0, 1\}$ are the adjacency matrix elements capturing the presence or absence of a link between nodes $i$ and $j$. The challenge resides in the fact that only a small fraction of all actual contacts are captured and recorded by our crowd-sourcing approach; reconstructed contact networks thus represent samples of the complete population network of interest where most nodes and links have been removed [3, 4]. We use an approach based on Horvitz-Thompson network sampling theory [3] and topological information from complete contact networks in the literature [5, 6, 7] to correct for sampling effects.

*S3.2. The sampling process induced by the data collection app*

The nature of our reconstructed contact networks as samples of the actual object of interest stems from several aspects inherent to our GPS crowd-sourcing method. Specifically, mobile device users install an app and opt in to the creation of samples including time and GPS location information, in response to certain triggers such as motion or activity on the phone. (i) Only a fraction of the population elects to install the app and are thus able to contribute (node sampling), thus giving rise to a panel of approximately 1 million users in Germany out of a population of 83 million. (ii) Participating individuals do not send samples continuously in time, but only occasionally upon trigger events (edge sampling). As a result, most real-world contact events go undetected.

The emerging mathematical picture of the network sampling process induced by the crowd-sourcing app is shown in Figure 1(b): An initial node sampling step retains only nodes which have the app installed on their device, which occurs with probability $p \approx 0.01$. Edges between retained nodes are also retained. Second, an additional edge sampling step retains only those edges where both involved devices create a sample during the contact; the simultaneity of samples (modulo 2 min) from different devices (Figure 1(c)) is a necessary condition for the recording of contacts and occurs with probability $q$.

The sampling parameters $p$ and $q$ are not constant, as app usage as well as trigger events for sample creation are subject to change over time. To achieve a persistent measurement of daily contact network statistics such as $CX$, the statistics obtained from network samples must be scaled by daily values of $p$ and $q$. Most importantly, $q$ is strongly affected by app updates, but previous modeling [8] is restricted to the node sampling part of the overall sampling process. In consequence, time series covering extended periods of time (3 years in this paper) represent a convolution of actual changes in contact networks in response to e.g. lockdowns and spurious changes inflicted by events unrelated to contact behaviour. Correcting these time series for app usage and software-related effects is key towards properly measuring actual epidemic factors.

Higher-order sampling effects, such as changes in phone usage in synchrony with contact behaviour changes as daily routines change, are not captured by our approach.

*S3.3. Horvitz-Thompson estimation of $CX$ from network samples*

Using Horvitz-Thompson network sampling theory, we here derive the relationship between the Contact Index $CX$ of the complete network $G$ and its counterpart $CX^*$ in a sample $G^*$ of this network obtained through the sampling process described above. Observables $\mathcal{O}$ ($\mathcal{O}^*$) in $G$ ($G^*$) are denoted without (with) an asterisk. The theory states that, for any edge observable $\mathcal{O}_{ij}$, we have that in expectation [3]

$$\sum_{ij\in G} \mathcal{O}_{ij} = \sum_{ij\in G^*} \frac{\mathcal{O}^*_{ij}}{\pi_{ij}}, \tag{S4}$$

where $\pi_{ij}$ is the probability of retaining the edge between nodes $i$ and $j$ upon sampling the original network.

For the moments $\langle k\rangle$ and $\langle k^2\rangle$ in the complete network, we thus find

$$\langle k\rangle = \frac{1}{N}\sum_{i\in G} k_i = \frac{1}{N}\sum_{ij\in G} a_{ij} = \frac{1}{N}\sum_{ij\in G^*} \frac{a^*_{ij}}{p_i p_j q_{ij}} \tag{S5}$$

and

$$\langle k^2\rangle = \frac{1}{N}\sum_{i\in G} k_i^2 = \frac{1}{N}\sum_{ij\ell\in G} a_{ij}a_{i\ell} = \frac{1}{N}\sum_{ij\neq\ell\in G^*} \frac{a^*_{ij}a^*_{i\ell}}{p_i p_j q_{ij} p_\ell q_{i\ell}} + \frac{1}{N}\sum_{ij\in G^*} \frac{a^*_{ij}}{p_i p_j q_{ij}}, \tag{S6}$$

where the last equality in each case applies Eq. (S4) to the adjacency matrix entries $\mathcal{O}_{ij} = a_{ij}$. The overall probability of retaining a single edge reads $\pi_{ij} = p_i p_j q_{ij}$ as used in Eq. (S5); it requires both retaining the nodes (with probabilities $p_i$ and $p_j$) and subsequently the edge itself (with probability $q_{ij}$). In Eq. (S6), we split the sum into terms where $j \neq \ell$ and $j = \ell$: In the former case, a second edge between $i$ and $\ell$ needs to be retained (with conditional probability $\pi_{i\ell} = p_\ell q_{i\ell}$) after a first edge between $i$ and $j$ has been retained (again with probability $\pi_{ij} = p_i p_j q_{ij}$).

Thus, assuming uniform sampling parameters $p_i \equiv p$ and $q_{ij} \equiv q$ across the network, which implies that the number of nodes scales as $N^* = pN$, we have

$$\langle k\rangle = \frac{1}{Np^2q}\sum_{i\in G^*} k_i^* = \frac{1}{pq}\frac{1}{N^*}\sum_{i\in G^*} k_i^* = \frac{\langle k^*\rangle^*}{pq} \tag{S7}$$

and

$$\begin{aligned}\langle k^2\rangle &= \frac{1}{Np^3q^2}\sum_{i\in G^*} k_i^*(k_i^*-1) + \frac{1}{Np^2q}\sum_{i\in G^*} k_i \\ &= \frac{1}{p^2q^2}\frac{1}{N^*}\sum_{i\in G^*} k_i^*(k_i^*-1) + \frac{1}{pq}\frac{1}{N^*}\sum_{i\in G^*} k_i \\ &= \frac{\langle k^{*2}\rangle^* - \langle k^*\rangle^*}{p^2q^2} + \frac{\langle k^*\rangle^*}{pq},\end{aligned} \tag{S8}$$

where $\langle\cdot\rangle^*$ denotes the average taken within the sample network. As a result, the Contact Index $CX$ is estimated as

$$CX = \frac{\langle k^2\rangle}{\langle k\rangle} = 1 + \frac{1}{pq}\left(\frac{\langle k^{*2}\rangle^*}{\langle k^*\rangle^*} - 1\right) = 1 + \frac{CX^* - 1}{pq}. \tag{S9}$$

Upon setting $q = 1$, we recover previous results [8] without the additional edge sampling step once the nodes have been sampled.

The node sampling probability $p = \frac{N^*}{N}$ is simply given by the population share participating in the crowd-sourcing and the probability of retaining non-unique contact links $q$ by the rate of simultaneous samples between pairs of nodes; both devices must create samples at the same time (modulo 2 min). Assuming that in the 2 min-interval $t$ ($t = 1, 2, \ldots, T = 720$ for a day of $720 \cdot 2\,\text{min} = 1440\,\text{min}$) $N^*(t) \leq N^*$ devices among the $N^*$ observed over the full day are active, we take

$$q = \left\langle \frac{N^*(t)(N^*(t)-1)}{N^*(N^*-1)} \right\rangle_t = \frac{1}{T}\sum_{t=1}^{T} \frac{N^*(t)(N^*(t)-1)}{N^*(N^*-1)}, \tag{S10}$$

i.e., the average fraction of device pairs with simultaneous pings among all possible pairs. Eq. (S10) uses the fact that at any given time $t$, devices have uncorrelated activity patterns, i.e. they create samples independently from one another (Figure 1(d)). Note, however, that there is a systematic variation in the number of active numbers $N^*(t)$ over the day, namely that devices are more active during daytime than at night (Figure 1(d)). By taking averages over the day, we expect to slightly underestimate the true $q$, as this collective daily activity pattern induces correlation between devices (Figure 1(d)).

In the case of unique contact networks relevant to our purposes, however, inferring original network properties from samples comes with its own intricacies because of structural information loss, as discussed hereafter.

*S3.4. Computing the edge sampling probability q for unique contacts*

Contact numbers in unique versus non-unique contact networks have different interpretations [9]: The non-unique case comprises all contacts, including repeated ones, regardless of how they are distributed across the network. In contrast, the unique case counts pairs connected by any number of contacts and, as such, focuses on the topological features of the network. Horvitz-Thompson theory appears to be limited to the non-unique counts: Its success in connecting contact counts between original and sample networks relies on the independent nature of edge sampling – each of $w_{ij}$ non-unique edges between nodes $i$ and $j$ is sampled independently with probability $q$. It fails for unique contacts because of a coupling effect: In order for a unique link to be retained upon sampling, at least one among $w_{ij} > 0$ non-unique links must be retained. However, we can rescue Horvitz-Thompson theory by considering independent survival of unique links with an effective edge sampling probability $q_{\text{eff}}$ (multilink density). Denoting by $w^*$ the remaining number of non-unique links after sampling ($0 \leq w^*_{ij} \leq w_{ij}$), this probability is given by

$$q_{\text{eff}} = P(w^* > 0 | w > 0) = \frac{P(w^* > 0)}{P(w > 0)}, \tag{S11}$$

where $P(w)$ defines a weight distribution in the network, i.e. the probability that a pair of nodes is connected by $w$ non-unique links.

The failure to infer original network properties from samples can be intuited by the destruction of structural network information upon edge sampling: Figure 1(f) illustrates how two networks with similar non-unique edge number but distinct topologies lead to similar sample networks upon edge sampling (green arrows). In consequence, the distinct original network topologies are indistinguishable from the sample as only source of information (red arrows) [10]. This topological information loss is also reflected by the non-injective relation between unique and non-unique weights, $a_{ij} = \text{sgn}(w_{ij})$; while the non-unique edge count $w_{ij} > 0$ indicates the presence of a unique link $a_{ij} > 0$, the reverse is not true.

Eq. (S11) reveals that $q_{\text{eff}}$ explicitly depends on unknown properties of the original network $P(w)$, which is where the missing structural information steps in. In consequence, the sampling parameter $q_{\text{eff}}$ is not fully determined by the crowd-sourcing data, but requires additional knowledge about structural features of population-wide contact networks. To fill the gap, we devise a Bayesian approach in combination with prior information from *complete* contact networks reported in the literature [5, 6, 7]. We observe a common shape of $P(w|w > 0)$ (Figure 1(g)) across a variety of contexts (cruiseship, university campus, small city), thus suggesting universal topological features in human contact networks also applicable as prior information to our case.

The cruiseship dataset provides the durations of all contact events between all pairs of individuals for 4 cruises of total duration of 37 h each, applying a 2 m proximity threshold to define a contact. We define the weight for a given pair as the (rounded) cumulative number of 2 min intervals spent in contact. The city dataset records all encounters within 50 m between any two individuals from a sample of 4 % of the city population on 3 consecutive days between 7am and 11pm, but not the duration of encounter. Similarly, the university dataset records all encounters between members of the freshmen class within $5 - 10$ m over the course of 28 days. While there is population sampling in the latter two datasets, the data is complete in a sense that within the population sample all contacts are systematically recorded (i.e., no edge sampling). For each day, we define the weights as the number of encounters between pairs of individuals.

More precisely, all weight distributions appear to be well fitted by zeta distributions (i.e. discrete power-law distributions), $P(w|w > 0) = w^{-(1+\alpha)}/\zeta(1 + \alpha)$, $\alpha > 0$ with exponents $\alpha$ inferred through maximum log-likelihood maximization using the log-likelihood function

$$\mathcal{L}(\alpha) = -\sum_{w>0} N(w) \left[\ln(\zeta(1 + \alpha)) + (1 + \alpha) \ln(w)\right], \tag{S12}$$

where $N(w)$ is the number of links in the network with weight $w$ and $\zeta(\cdot)$ is the Riemann zeta function. The equation $\frac{\partial \mathcal{L}}{\partial \alpha}(\hat{\alpha}) = 0$ then has approximate solution [11]

$$\hat{\alpha} \approx \left( \frac{\sum_{w>0} N(w) \ln(w)}{\sum_{w>0} N(w)} + \ln(2) \right)^{-1}. \tag{S13}$$

The distribution of values for $\alpha$ across all daily networks is shown in the inset of Figure 1(g). Note that we do not assert by our analysis that power laws are the true mechanism behind the observed networks [11, 10, 12]. Rather, we will use this model and its topological information as an approximate representation of the observed networks to perform the normalization of the sampling parameter $q$. More precisely, we will use a discrete power-law prior distribution $P_0(w|w > 0)$ in the following. Also note that $P_0(w = 0)$ is excluded because the density of the network $(1 - P_0(w))$ is expected to be vastly different between spatially confined and spatially extended contact networks.

To express $q_{\text{eff}}$ in terms of $q$ and the prior distribution $P_0(w)$, we use the binomial distribution connecting $w$ and $w^*$,

$$P(w^*|w) = \binom{w}{w^*} q^{w^*} (1 - q)^{w-w^*}, \tag{S14}$$

as well as a Bayesian update equation for $P(w)$,

$$P(w) = \sum_{w^* \geq 0} P(w|w^*)P(w^*) = \sum_{w^* \geq 0} \frac{P(w^*|w)P_0(w)}{P_0(w^*)} P(w^*), \tag{S15}$$

where the second equality makes use of the Bayesian theorem $P(w^*|w)P(w) = P(w|w^*)P(w^*)$. Evaluating the update equation at $w = 0$ and using $P(w = 0|w^*) = \delta_{w^*0}$ as per the binomial distribution shows that

$$P(w = 0)P_0(w^* = 0) = P_0(w = 0)P(w^* = 0). \tag{S16}$$

Because of the following series of equalities,

$$\begin{aligned}
P(w > 0)P_0(w^* > 0) &= (1 - P(w = 0))(1 - P_0(w^* = 0)) \\
&= 1 - P(w = 0) - P_0(w^* = 0) + P(w = 0)P_0(w^* = 0) \\
&= 1 - P(w = 0) - P_0(w^* = 0) + P_0(w = 0)P(w^* = 0) \\
&= 1 - P_0(w = 0) - P(w^* = 0) + P_0(w = 0)P(w^* = 0) \\
&\qquad + [P_0(w = 0) - P(w = 0)] + [P(w^* = 0) - P_0(w^* = 0)] \\
&= (1 - P_0(w = 0))(1 - P(w^* = 0)) \\
&\qquad + [P_0(w = 0) - P(w = 0)] + [P(w^* = 0) - P_0(w^* = 0)] \\
&= P_0(w > 0)P(w^* > 0) \\
&\qquad + [P_0(w = 0) - P(w = 0)] + [P(w^* = 0) - P_0(w^* = 0)],
\end{aligned} \tag{S17}$$

where the second equality makes use of Eq. (S16), we imply that

$$\begin{aligned}
P(w > 0)P_0(w^* > 0) = P_0(w > 0)P(w^* > 0) &+ [P_0(w = 0) - P(w = 0)] \\
&+ [P(w^* = 0) - P_0(w^* = 0)].
\end{aligned} \tag{S18}$$

Thus, under the assumption that the prior and actual networks are similarly dense, i.e. $|P(w^{(*)} = 0) - P_0(w^{(*)} = 0)| \ll 1$, we can neglect the terms in brackets $[\dots]$ in Eq. (S18) to obtain

$$\begin{aligned}
q_{\text{eff}} &= \frac{P(w^* > 0)}{P(w > 0)} \approx \frac{P_0(w^* > 0)}{P_0(w > 0)} = \frac{1 - P_0(w^* = 0)}{P_0(w > 0)} = \sum_{w>0} \frac{P_0(w)}{P_0(w > 0)} \left[1 - (1 - q)^w\right] \\
&= \sum_{w>0} P_0(w|w > 0) \left[1 - (1 - q)^w\right] = \sum_{w>0} \frac{w^{-(1+\alpha)}}{\zeta(1 + \alpha)} [1 - (1 - q)^w] = 1 - G(1 - q),
\end{aligned} \tag{S19}$$

where the approximation uses Eq. (S18), the second equality uses the binomial distribution, the third equality uses the definition of conditional probabilities, and the last equality uses the zeta distribution as a model for $P_0(w|w > 0)$. Moreover, $G(\xi) = \sum_{w>0} \frac{\xi^{-(1+\alpha)}}{\zeta(1+\alpha)} \xi^w = \zeta(1 + \alpha)^{-1} \mathrm{Li}_{1+\alpha}(\xi)$ is the probability generating function of the zeta distribution and $\mathrm{Li}_{\cdot}(\cdot)$ is the polylogarithm. Expectedly, $q_{\text{eff}}$ is a non-linear and strictly monotonously increasing function of $q$.

### S4. Spatial heterogeneity

#### *S4.1. Correcting for spatially heterogeneous sampling: the role of soccer stadiums*

We found that GPS location data at mass events in certain, at least partially roofed locations can be flawed. This applies in particular to soccer matches in large stadiums: Some crowd-sourcing samples are clustered in specific areas within stadiums (Figure S3(a)) which appears implausible. This spurious co-location of devices leads to false contacts which need to be identified and removed from the Contact Index analyses.

The Android operating system uses 3 different technologies to determine device locations: pure GPS (samples of type `GPS`) as well as GPS in combination with two Android-specific methods (samples of type `NET` or `FUSED`). Using GPS location data labelled with ground truth locations from an on-site experiment at the Olympiastadion Berlin, we revealed that samples of type `GPS` are reliable, while certain stadium areas appear to be attractors for many samples of type `NET` and `FUSED` (Figure S3(b)). Therefore, we decide to remove contacts occurring in these apparent clusters, which is achieved through retaining all contact pairs where at least one (of two) co-located samples is of type `GPS`. All stadiums connected to soccer teams in the first 3 national soccer leagues receive this special treatment.

Only about 10 % of in-stadium samples are of type `GPS`. Having thus a 10-fold less dense sampling inside of stadiums as compared to outside of stadiums has an impact on the effective sampling parameters $p$ and in particular $q_{\text{eff}}$ inside of stadiums. This leads us to a situation of heterogeneous sampling within the contact network, with some network portions being sampled differently as compared to others. Indexing the two geographically distinct regions "outside stadium" and "inside stadium" by 1 and 2, respectively, we identify the region-specific sampling parameters as

$$p_1 = \frac{N_1^*}{N_1}, \qquad p_2 = \frac{N_2^*}{N_2} \tag{S20}$$

and

$$q_1 = \left\langle \frac{N_1^*(t)(N_1^*(t)-1)}{N_1(N_1-1)} \right\rangle_t, \qquad q_2 = \left\langle \frac{N_2^*(t)(N_2^*(t)-1)}{N_2(N_2-1)} \right\rangle_t, \tag{S21}$$

where $N_{1/2}^*$ and $N_{1/2}$ are respectively the number of distinct observed devices and the total population of regions 1 and 2. For the sake of convenience, we use $N_1^* \approx N^*$ and $N_1 \approx N$, which reflects that devices detected inside stadiums are likely to be also detected outside of stadiums on the same day (equivalently: we ignore the number of devices exclusively found inside stadiums). As such, $p_1$ and $q_1$ are again equivalent to $p$ and $q$ under uniform sampling.

Computing $p_2$ requires knowledge of $N_2$, i.e. the stadium population present during a match. For regularly scheduled matches in the 3 national soccer leagues, among others, this information can be readily gathered from [13]. We find that the node sampling inside of relevant stadiums during soccer matches after removal of samples of type `NET` or `FUSED` fluctuates around 73.0 % of the level observed outside of stadiums, i.e. $p_2 = 0.730 p_1$. To simplify the analysis and take into account irregular soccer events such as international matches, we fix $p_2$ to 73 % of $p_1$ in all stadiums at all times, thus neglecting day- or event-specific variations in the value of $p_2$.

To define the overall Contact Index in this non-uniform sampling case, we need to take a different perspective on its definition $CX = \frac{\langle k^2 \rangle}{\langle k \rangle}$. Specifically, $\langle k \rangle$ and $\langle k^2 \rangle$ count 1-step and 2-step paths along links in the contact network. In the heterogeneously sampled case, we need to count 1-step and 2-step paths within as well as across regions. To this aim, we partition the overall unique contact network into region-specific unique contact networks $G_1$ and $G_2$ with adjacency matrices $\mathbf{A}_1$ and $\mathbf{A}_2$ such that $a_{1,ij} = 1$ if $i$ and $j$ are in contact in region 1 and similarly for region 2. Note that $a_{1,ij} = a_{2,ij} = 1$ for any pair that is in contact both inside and outside of stadiums. Within regions, where sampling is again uniform, the estimates of 1-step path counts $K_{1/2}$ and 2-step path counts $K_{11/22}$ are estimated following Eqs. (S7) and (S8) as

$$\begin{aligned} K_{1/2} &= \frac{1}{p_{1/2}^2 q_{1/2}} \sum_{ij \in G^*} a^*_{1/2,ij}, \\ K_{11/22} &= \frac{1}{p_{1/2}^3 q_{1/2}^2} \left( \sum_{ij \neq \ell \in G^*} a^*_{1/2,ij} a^*_{1/2,i\ell} - (1 - p_{1/2} q_{1/2}) \sum_{ij \in G^*} a^*_{1/2,ij} \right). \end{aligned} \tag{S22}$$

Moreover, 2-step paths can span across regions with, for instance, the center node $i$ having contact with $j$ outside the stadium and with $\ell$ inside the stadium. Such paths would be retained under network sampling with proba $p_{1\&2,i} p_{1,j} p_{2,\ell} q_{1,ij} q_{2,i\ell}$, where $p_{1\&2}$ is the fraction of nodes found both in region 1 and 2. We approximate $p_{1\&2} \approx p_2$ to reflect that devices detected inside stadiums are likely to also be detected outside stadiums ($p_{1|2} = 1$) on a given day. The Horvitz-Thompson estimation for the number of such paths $(K_{12} + K_{21}) = 2K_{12}$ thus reads

$$K_{12} + K_{21} = \frac{2}{p_1 p_2 p_{1\&2} q_1 q_2} \sum_{ij\ell \in G^*} a^*_{1,ij} a^*_{2,i\ell}. \tag{S23}$$

Overall, the contact number moments then read

$$\langle k \rangle = \frac{K_1 + K_2}{N} = \frac{1}{\frac{N_1^*}{p_1} + \frac{N_2^*}{p_2}} \left( \frac{1}{p_1^2 q_1} \sum_{ij \in G^*} a^*_{1,ij} + \frac{1}{p_2^2 q_2} \sum_{ij \in G^*} a^*_{2,ij} \right) \tag{S24}$$

and

$$\begin{aligned} \langle k^2 \rangle &= \frac{K_{11} + K_{22} + K_{12} + K_{21}}{N} \\ &= \frac{1}{\frac{N_1^*}{p_1} + \frac{N_2^*}{p_2}} \left[ \frac{1}{p_1^3 q_1^2} \left( \sum_{ij \neq \ell \in G^*} a^*_{1,ij} a^*_{1,i\ell} - (1 - p_1 q_1) \sum_{ij \in G^*} a^*_{1,ij} \right) \right. \quad \text{(S25)} \\ &\quad + \frac{1}{p_2^3 q_2^2} \left( \sum_{ij \neq \ell \in G^*} a^*_{2,ij} a^*_{2,i\ell} - (1 - p_2 q_2) \sum_{ij \in G^*} a^*_{2,ij} \right) \\ &\quad \left. + \frac{2}{p_1 p_2 p_{1\&2} q_1 q_2} \sum_{ij\ell \in G^*} a^*_{1,ij} a^*_{2,i\ell} \right]. \quad \text{(S26)} \end{aligned}$$

Note that contacts occurring in both regions (such as friends watching a match together and then going to a restaurant together) are counted twice, once in each region. In the case of non-unique contacts where repeated contacts should not be counted, we should therefore introduce a cross-region correction term $\left(-\sum_{ij} a_{1,ij}a_{2,ij}\right)$ for $\langle k\rangle$ and $\left(-\sum_{ij\ell} a_{1,ij}(a_{1,i\ell}a_{2,i\ell}) - \sum_{ij\ell}(a_{1,ij}a_{2,ij})a_{2,i\ell}\right)$ for $\langle k^2\rangle$ to remove multiple counts of contacts present in both regions. For simplicity, we here neglect these corrections, as such contacts are rare. Figure S3(c) compares the overall Contact Index $CX$ and contributions from stadiums $\frac{K_{22}}{K_1+K_2}$: Expectedly, stadium contributions are indistinguishable from zero during Christmas holidays and lockdown periods. Stadium contributions are small (by a factor of at least $\sim 10$) at all times compared to overall contact levels [14].

*S4.2. Spatially heterogeneous contact patterns*

To demonstrate the dimensionality and comparability of the Contact Index $CX$, we compute $CX$ separately for all 16 German federal states: We partition the daily Germany-wide contact network by coloring the nodes according to their home states. The home state of a device is inferred on a monthly basis as the largest spatial cluster of samples among all of its samples over the course of the month. For any federal state, $CX = \frac{\langle k^2\rangle}{\langle k\rangle}$ is then computed from the distribution of unique contact numbers $k_i$ among all observed devices $i$ based in that state, including cross-state contacts. State-level sampling parameters $p$ and $q_{\text{eff}}$ are computed in complete analogy to the national level.

The classification of nodes by home location is relevant for epidemic statistics, as infection test results are typically recorded and associated with an individual's home location. In 2020, high daily $CX$ values at the state level are indicative of high state-specific 7-day average SARS-CoV-2 reproduction numbers $R_{\text{eff}}$, computed from state-level infection numbers, about $\Delta t = 16$ days later (Figure S3(e)). Yet, increasing the spatial resolution for $CX$ or, equivalently, computing $CX$ for smaller portions of the network is limited by the sampling depth of our crowd-sourcing approach: The signal-to-noise ratio is decreased as fewer absolute numbers of individuals are available to estimate the moments $\langle k\rangle$ and in particular $\langle k^2\rangle$. Upon estimating confidence intervals for $CX$, we found that nation- and state-level $CX$ are significant, but most county-level $CX$ values are not.

Comparing long-term averaged $CX$ values between federal states reflects the expectation that levels of contacts tend to be globally higher in city states with high population density (Berlin, Hamburg, Bremen) than in geographically wide-stretching states and also higher in East German states compared to West German states (Figure S3(d)).

### S5. Determining and analyzing relative transmissibility $\langle T\rangle$ using $CX$

*S5.1. Calibration of $R$ and $CX$ using 2020-specific data*

Throughout the manuscript, we use centered 7-day averages $\langle CX\rangle(t) = \frac{1}{7}\sum_{\tau=-3}^{3} CX(t+\tau)$ to eliminate weekly periodicity in $CX$ and maintain only its long-term trend, but skip the brackets $\langle\cdot\rangle$ for clarity. We use daily Contact Index values $CX$ and SARS-CoV-2 reproduction numbers $R_{\text{eff}}$ (now-cast data recorded by RKI [15]) in the time window between 04/01/2020 and 12/31/2020 to establish the relationship between contact and transmission levels in absence of other factors: SARS-CoV-2 testing has become widely accessible by April 2020, while the turn of the year 2020/2021 marks the beginning of vaccine campaigns and the takeover of immune escape variants other than wild-type SARS-CoV-2. This time range covers parts of the first and second lockdowns in Germany as well as the comparatively unregulated summer 2020, thus providing ample amount of dynamics in terms of contact behaviour (Figure S1(a)) to study the correlation between the $CX$ and $R_{\text{eff}}$ time series.

For $\Delta t = 16$ days, time lead for which the linear Pearson correlation $\text{Corr}[CX, R_{\text{eff}}](\Delta t)$ between $CX$ and $R_{\text{eff}}$ is maximal (Figure S1(a, right inset)), we perform a linear regression of the data [8],

$$R_{\text{WT}}(t + \Delta t) = R_{\text{WT}}(CX(t)) = a + b \cdot CX(t), \tag{S27}$$

with parameters $a$ and $b$ found by minimizing the unweighted sum of squared residuals to be $a = 0.56$ and $b = 0.01$ (Figure S1(a, left inset)). The time lead of $\Delta t = 16$ days between the contact and the day appointed by the RKI for now-cast $R_{\text{eff}}$ values is explained, among other things, by the incubation period, delay in reporting, and averaging intervals.

*S5.2. Relative transmissibility: overall dynamics less the contacts*

Given the value of $CX$ on day $t$, Eq. (S27) provides a prediction of $R_{\text{WT}}$ on day $t + \Delta t$. Then, we can interpret the discrepancy (ratio) between the prediction $R_{\text{WT}}$ and official $R_{\text{eff}}$ value

$$T(t) = \frac{R_{\text{eff}}(t + \Delta t)}{R_{\text{WT}}(CX(t))}. \tag{S28}$$

as the discrepancy between wild-type transmission efficiency under unperturbed conditions and actual transmission efficiency, i.e. the *relative transmissibility* of the contagion. This leads to a noisy time series $T(t)$ for the slowly varying relative transmissibility whose trend is, by the easiest of all methods, captured by a smoothened signal $\langle T \rangle(t) = \frac{1}{2\tau+1} \sum_{\Delta t=-\tau}^{\tau} T(t)$ where we here used $\tau = 30$ days, i.e. sliding centered averages over 2 months.

*S5.3. Confounding: correlation of $\langle T \rangle$ with other time series*

To interpret the quantity $\langle T \rangle(t)$, we study the correlation between its daily trends ($\langle T \rangle(t) - \langle T \rangle(t-1)$) with those of various other time series since the end of the time window used for calibration of $CX$ and $R$ (2021/01/01). Results are shown in Figure S1(b,c). We here include epidemic factors (virus mutant frequencies [16], population share per vaccination status [17]), test positivity [18], averages of locally measured prevalence [19]), and network sampling ($p$ and $q$), as well as other topological features of the measured contact networks (cluster coefficient, smallworldness, etc.). Similarly to $\langle T \rangle(t)$, we first compute a temporal average $\langle \mathcal{O} \rangle(t) = \frac{1}{2\tau+1} \sum_{\Delta t=-\tau}^{\tau} \mathcal{O}(t)$ with $\tau = 30$ days for any quantity $\mathcal{O}$ before computing Pearson correlations between $\langle T \rangle(t)$ and $\langle \mathcal{O} \rangle(t)$.

To correlate $\langle T \rangle(t)$ with SARS-CoV-2 evolutionary dynamics, we define a time series given by

$$\mathcal{O}(t) = \sum_{\mu} |f_\mu(t) - f_\mu(t-1)| \tag{S29}$$

where $f_\mu(t)$ denotes the centered 7-day average frequency of SARS-CoV-2 mutant $\mu$ on day $t$. This time series peaks whenever absolute frequency slopes are high, i.e. whenever a takeover by a new variant occurs.

SARS-CoV-2 prevalence studies are restricted in space and time: Typically, blood samples are taken and analyzed at the city or county level over the range of a few weeks. For every day $t$, we here use the average prevalence value across prevalence studies at different locations covering that day $t$, if any, as an indicator for the Germany-wide SARS-CoV-2 prevalence $\mathcal{O}(t)$.

Unlike for the Contact Index $CX$ which is tractable through Horvitz-Thompson theory, the scaling of topological features upon network sampling is more intricate or even impossible. Network sampling was shown to affect different topology metrics in various ways [20, 21]. Here, we simply use topological features of the measured sample networks. However, since the sampling scheme remains itself overall unchanged, we expect potential biases to be constant in time and trends in sample networks to reflect actual trends within the underlying complete networks. For every day $t$, we aggregate the measured networks between $t$ and $t+6$ to increase the statistical basis for the computation of topology measures, i.e. we include a link between a given pair of devices if there is a link on at least one day of the 7 days between $t$ and $t+6$.

### S6. Epidemic forecast

#### *S6.1. SARS-CoV-2*

The challenge of epidemic forecast consists in the accurate prediction of current and future infection numbers or reproduction numbers. Now-cast $R_{\text{eff}}$ values, as published by the RKI for SARS-CoV-2 [15], do not provide a real-time picture of the infection dynamics, as they reflect past infections arising from past contacts with a delay of around $\Delta t = 16$ days. As a result, denoting the current day by $t_0$, infection surveillance can provide insights only up to day $t_0 - \Delta t$, i.e. up to $2-3$ weeks ago.

On the contrary, our crowd-sourcing and $CX$ data is being collected and processed in near real-time. Currently, the data import process from mobile devices induces a delay of only 2 days, but which is being further reduced via optimization of the data pipeline. The real-time nature of $CX$ thus leads to a straight-forward forecast of recent and current reproduction numbers up to $t_0$, under the assumption of unchanged relative transmissibility trend. The relative transmissibility $\langle T \rangle$ itself inherits its delay of $\Delta t = 16$ days from $R_{\text{eff}}$ and is projected beyond $t_0 - \Delta t$ (see below). Beyond $t_0$, both $CX$ and relative transmissibility $T$ need to be projected from previous data.

For given $t_0$, we fit auto-regressive integrated moving-average (ARIMA) models to the time series: For $CX$, we use the last 60 data points up to $t_0$ to fit a model with auto-regressive order $p = 2$, differencing degree $d = 1$ and moving-average order $q = 2$. For $\langle T \rangle$, we use the last 180 data points up to $t_0 - \Delta t$ and $p = 2$, $d = 1$ and $q = 3$. We use the fitted models to project the time series up to 30 days into the future, i.e. up to $t_0 + 30$. The reproduction number forecast is then obtained from the $CX$ and $T$ time series via Eq. (4) and the $1\sigma$ (68 %) confidence intervals from the ARIMA models $\Delta CX$ and $\Delta T$ are propagated to $R_{\text{pred}}$ through

$$\Delta R_{\text{pred}} = (a + b \cdot CX)\Delta T + b \cdot \Delta CX \cdot T. \tag{S30}$$

As a null forecast that makes no use of our contact measurement, we project the infection surveillance data beyond $t_0 - \Delta t$ by fitting a model with $p = 0$, $d = 2$ and $q = 0$ to the last 60 days of $R_{\text{eff}}$ data.

To showcase and evaluate our epidemic forecast, we iterate $t_0$ between 10/01/2020 and 12/20/2022 and (i) compare $R_{\text{pred}}$ and its confidence interval with the actual $R_{\text{true}}$ for selected $t_0$ (Figure 3(a, upper panel)) and (ii) compare the distribution of residuals ($R_{\text{pred}} - R_{\text{true}}$) over all choices of $t_0$ at all time points between $t_0 - \Delta t$ and $t_0 + 30$ between the null and actual forecasts (Figure 3(a, lower panel)).

*S6.2. Influenza*

To demonstrate the broad applicability of our method to airborne transmissible diseases, we perform a forecast of Influenza infection levels equivalently to our SARS-CoV-2 forecast (Figure S2). The case of Influenza comes with two major limitations unrelated to our method: 1) Infection surveillance is not as systematic as for SARS-CoV-2. For Germany, the RKI publishes weekly infection numbers [22], from which we define a rough estimation of $R(t)$ as the ratio of the current $(t)$ and one-week prior $(t-7)$ smoothened infection numbers. Of note, the goal of this approach is solely to define a time series that represents the trends in infection levels, not to rigorously define reproduction numbers. 2) Unlike epidemic SARS-CoV-2, endemic Influenza has no phase with constant mutant background required for the calibration of $R$ and $CX$. For simplicity, we therefore assume a similar relationship as for SARS-CoV-2 and use identical regression parameters $a$ and $b$. This assumption, however, should only affect the scale of the quantities, not their trends and forecast performances.

### References

[1] N. Pitsianis, D. Floros, A.-S. Iliopoulos, X. Sun, Sg-t-sne-$\pi$: Swift neighbor embedding of sparse stochastic graphs, Journal of Open Source Software 4 (39) (2019) 1577.

[2] N. Pitsianis, A.-S. Iliopoulos, D. Floros, X. Sun, Spaceland embedding of sparse stochastic graphs, in: 2019 IEEE High Performance Extreme Computing Conference (HPEC), 2019, pp. 1–8. doi:10.1109/HPEC.2019.8916505.

[3] E. D. Kolaczyk, Statistical Analysis of Network Data, Springer New York, NY, 2009. doi:https://doi.org/10.1007/978-0-387-88146-1.

[4] P. Hu, W. C. Lau, A survey and taxonomy of graph sampling, CoRR abs/1308.5865 (2013). arXiv:1308.5865.
URL http://arxiv.org/abs/1308.5865

[5] R. Pung, J. A. Firth, L. G. Spurgin, V. J. Lee, A. J. Kucharski, Using high-resolution contact networks to evaluate sars-cov-2 transmission and control in large-scale multi-day events, Nature communications 13 (1) (2022) 1–11.

[6] P. Sapiezynski, A. Stopczynski, D. D. Lassen, S. Lehmann, Interaction data from the copenhagen networks study, Scientific Data 6 (1) (2019) 315.

[7] S. M. Kissler, P. Klepac, M. Tang, A. J. Conlan, J. R. Gog, Sparking “the bbc four pandemic”: Leveraging citizen science and mobile phones to model the spread of disease, bioRxiv (2020). arXiv:https://www.biorxiv.org/content/early/2020/05/12/479154.full.pdf, doi:10.1101/479154.
URL https://www.biorxiv.org/content/early/2020/05/12/479154

[8] S. Rüdiger, S. Konigorski, A. Rakowski, J. A. Edelman, D. Zernick, A. Thieme, C. Lippert, Predicting the sars-cov-2 effective reproduction number using bulk contact data from mobile phones, Proceedings of the National Academy of Sciences 118 (31) (2021) e2026731118. arXiv:https://www.pnas.org/doi/pdf/10.1073/pnas.2026731118, doi:10.1073/pnas.2026731118.
URL https://www.pnas.org/doi/abs/10.1073/pnas.2026731118

[9] M. A. Serrano, M. Boguñá, R. Pastor-Satorras, Correlations in weighted networks, Phys. Rev. E 74 (2006) 055101. doi:10.1103/PhysRevE.74.055101.
URL https://link.aps.org/doi/10.1103/PhysRevE.74.055101

[10] J.-D. J. Han, D. Dupuy, N. Bertin, M. E. Cusick, M. Vidal, Effect of sampling on topology predictions of protein-protein interaction networks, Nature biotechnology 23 (7) (2005) 839–844.

[11] A. Clauset, C. R. Shalizi, M. E. J. Newman, Power-law distributions in empirical data, SIAM Review 51 (4) (2009) 661–703. arXiv:https://doi.org/10.1137/070710111, doi:10.1137/070710111.
URL https://doi.org/10.1137/070710111

[12] R. Perline, Strong, weak and false inverse power laws, Statistical Science 20 (1) (2005) 68–88.
URL http://www.jstor.org/stable/20061161

[13] kicker.de, Kicker (2022).
URL https://www.kicker.de/

[14] B. García Bulle, D. Shen, D. Shah, A. E. Hosoi, Public health implications of opening national football league stadiums during the covid-19 pandemic, Proceedings of the National Academy of Sciences 119 (14) (2022) e2114226119.

[15] M. an der Heiden, Sars-cov-2-nowcasting und -r-schaetzung (Jan. 2023). doi:10.5281/zenodo.7571376.
URL https://doi.org/10.5281/zenodo.7571376

[16] R. Koch-Institut, Anzahl und anteile von voc und voi in deutschland (2023).
URL https://www.rki.de/DE/Content/InfAZ/N/Neuartiges_Coronavirus/Daten/VOC_VOI_Tabelle.xlsx?__blob=publicationFile

[17] R. Koch-Institut, Tabelle mit den gemeldeten impfungen nach bundesländern und impfquoten nach altersgruppen (2023).
URL https://www.rki.de/DE/Content/InfAZ/N/Neuartiges_Coronavirus/Daten/Impfquotenmonitoring.xlsx?__blob=publicationFile

[18] E. Mathieu, H. Ritchie, L. Rodés-Guirao, C. Appel, C. Giattino, J. Hasell, B. Macdonald, S. Dattani, D. Beltekian, E. Ortiz-Ospina, M. Roser, Coronavirus pandemic (covid-19) (2020).
URL https://ourworldindata.org/coronavirus

[19] H. Neuhauser, N. Buttmann-Schweiger, J. Fiebig, C. Poethko-Müller, F. Prütz, G. Sarganas Margolis, R. Thamm, M. Zimmermann, Observatorium serologischer Studien zu SARS-CoV-2 in Deutschland (Sep. 2022). doi:10.5281/zenodo.7043025.
URL https://doi.org/10.5281/zenodo.7043025

[20] J. D. Noh, Percolation transition in networks with degree-degree correlation, Phys. Rev. E 76 (2007) 026116. doi:10.1103/PhysRevE.76.026116.
URL https://link.aps.org/doi/10.1103/PhysRevE.76.026116

[21] S. H. Lee, P.-J. Kim, H. Jeong, Statistical properties of sampled networks, Phys. Rev. E 73 (2006) 016102. doi:10.1103/PhysRevE.73.016102.
URL https://link.aps.org/doi/10.1103/PhysRevE.73.016102

[22] R. Koch-Institut, Survstat@rki 2.0 (2023).
URL https://survstat.rki.de/

[23] T. Hale, N. Angrist, R. Goldszmidt, B. Kira, A. Petherick, T. Phillips, S. Webster, E. Cameron-Blake, L. Hallas, S. Majumdar, et al., A global panel database of pandemic policies (oxford covid-19 government response tracker), Nature human behaviour 5 (4) (2021) 529–538.

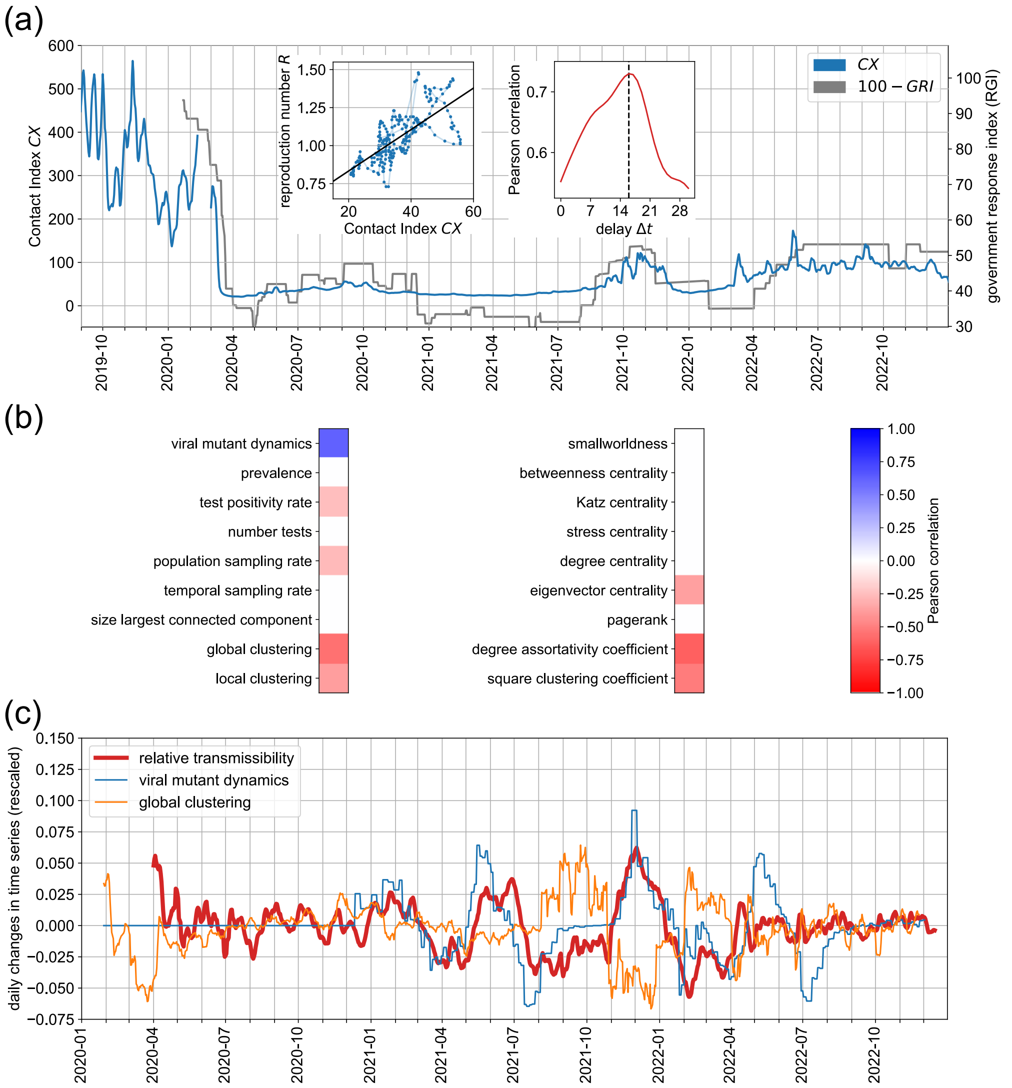

Figure S1: **(a)** Comparison of the Contact Index $CX$ (same as in Figure 2(a)) with the Government Response Index [23], indicating concurrent trends albeit no causal link between non-pharmaceutical health policies (NPIs) and contact levels. **(a, left inset)** Calibration of the Contact Index $CX(t)$ and SARS-CoV-2 effective reproduction numbers $R_{\mathrm{eff}}(t + \Delta t)$ ($\Delta t = 16$ days), independently recorded by the RKI [15], between April and December 2020 by linear regression. A linear relationship between $CX$ and $R_{\mathrm{eff}}$, with a certain temporal shift $\Delta t$ due to incubation time and testing/reporting delays, is expected in predominantly contact-driven epidemic trends (absence of immune escape variants and vaccination). **(a, right inset)** Pearson correlation between $CX(t)$ and $R_{\mathrm{eff}}(t + \Delta t)$ between April and December 2020 as a function of the time lead $\Delta t$. The correlation is highest for a time lead of $\Delta t = 16$ days, thus implying that $CX$ precedes reproduction numbers by about $2 - 3$ weeks. **(b)** Interpretation of relative transmissibility $\langle T \rangle(t)$: Correlation between SARS-CoV-2 relative transmissibility changes $\langle T \rangle(t) - \langle T \rangle(t-1)$ and various time series (frequency trends of variants, average of local prevalence levels, test positivity, network sampling parameters, and topological features of the contact network as estimated from network samples disregarding potential effects from network sampling). **(c)** Comparison of normalized time series between transmission trends and the most strongly correlated features identified in (b).

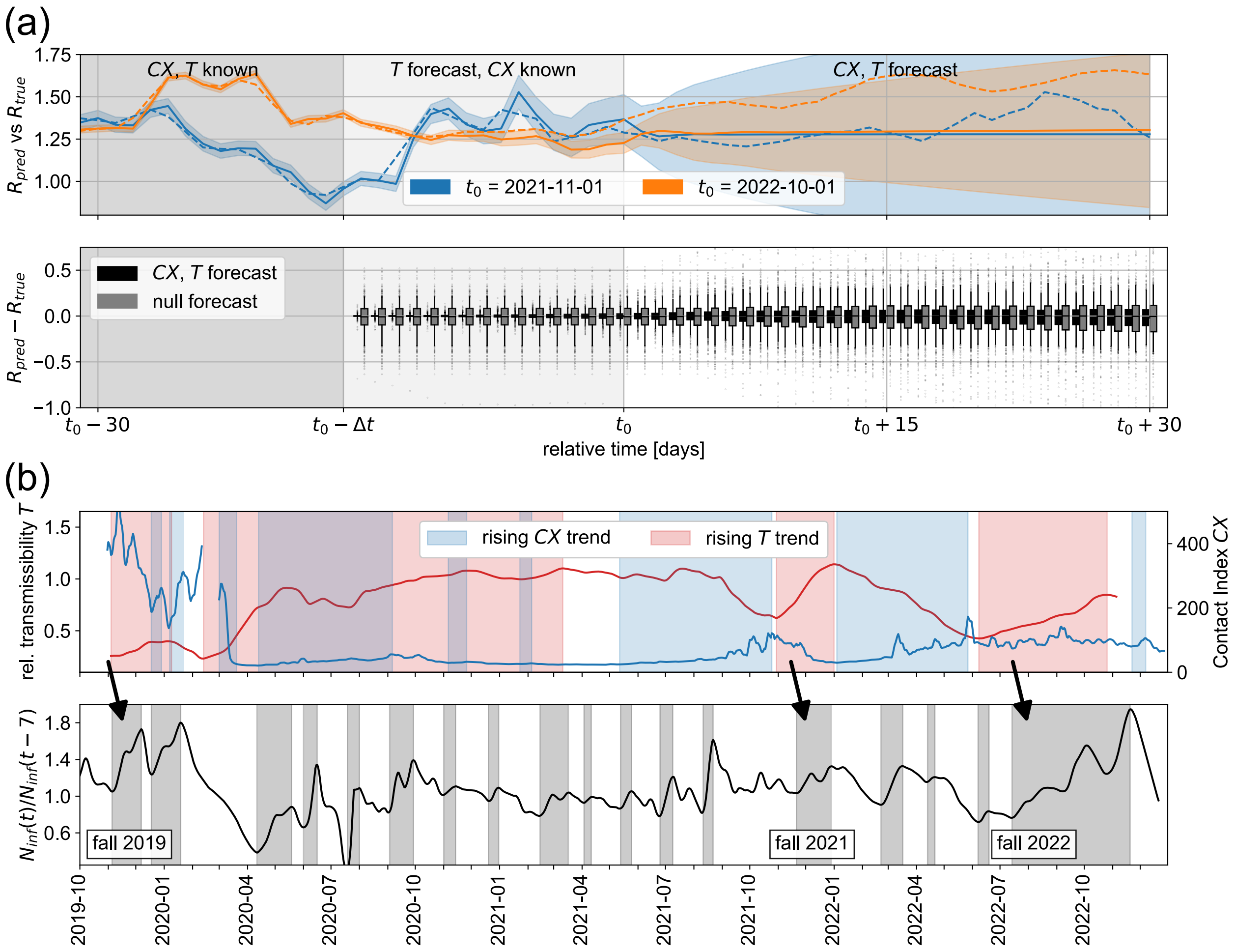

Figure S2: **(a)** Same as Figure 3(a), but for Influenza. **(b)** Same as Figure 3(b), but for Influenza.

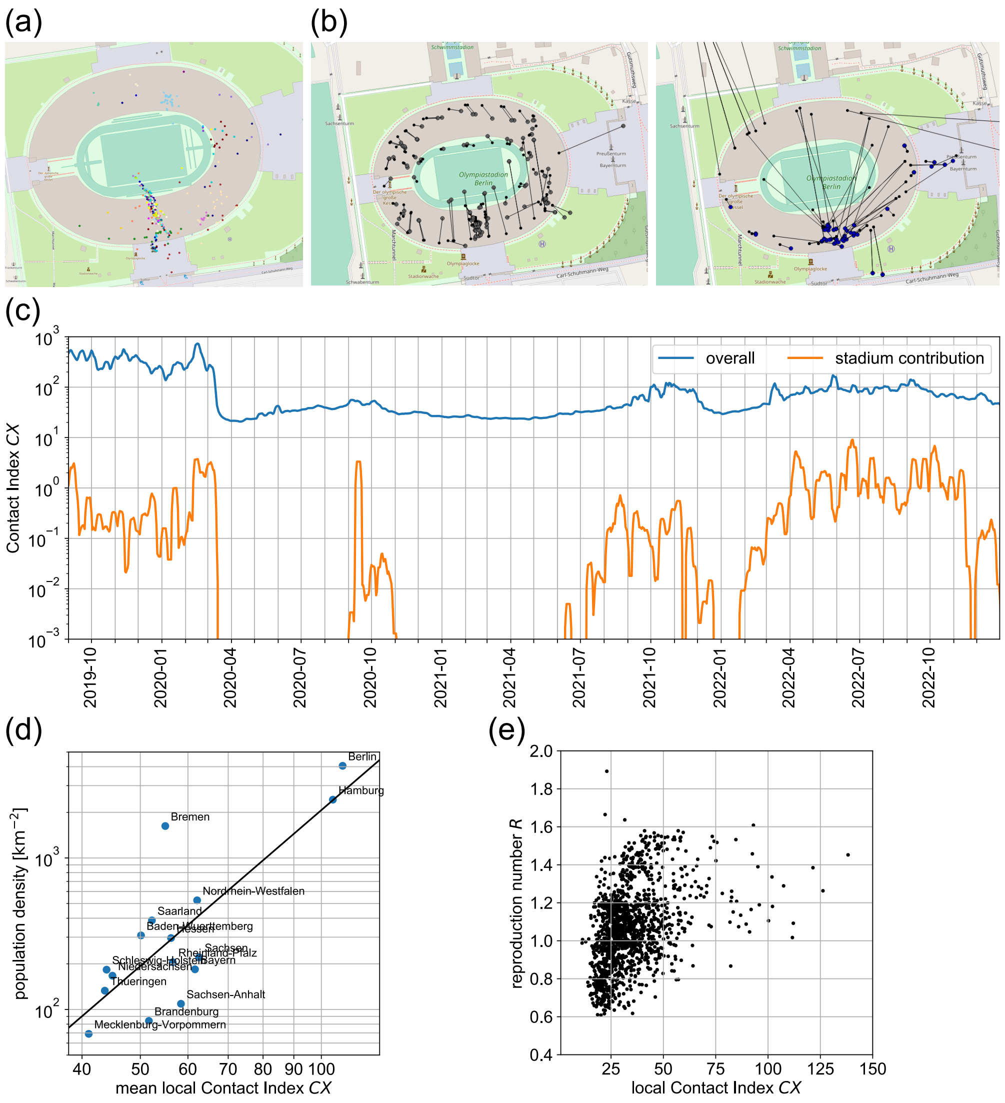

Figure S3: **Spatial heterogeneity of sampling and contact levels.** **(a)** Distribution of samples in the Olympiastadion Berlin area during the Berlin-Wolfsburg soccer match on 08/14/2021, showing an implausible concentration of samples in the southern part. **(b)** Comparison between positions determined by the app and actual, ground truth positions from an experiment conducted in the Olympiastadion Berlin. `GPS`-sourced locations (left) reflect true positions, while `NET`-sourced locations oftentimes are systematically off (right), with particular locations acting as attractors. (`GPS` and `NET` refer to distinct localization methods defined by the Android operating system.) **(c)** Comparison between overall $CX$ (same as Figure 2(a)) and its contributions from major soccer stadiums. Stadium attendance appears to have negligible impact on overall contact levels; note the log scale on vertical axis. Periods with stadium contribution below $10^{-3}$ are those where mass events were banned by health policy measures. **(d)** Relationship between average $CX$ values specific to federal states of Germany and their population densities. City states (Berlin, Hamburg) with the exception of Bremen expectedly tend towards higher $CX$ values. Eastern states tend towards higher $CX$ than Western states with similar population density. **(e)** Relationship between state-specific $CX$ and state-specific $R_{\text{eff}}$ values in 2020.
